# Cholinergic and lipid mediators crosstalk in Covid-19 and the impact of glucocorticoid therapy

**DOI:** 10.1101/2021.01.07.20248970

**Authors:** Malena M. Pérez, Vinícius E. Pimentel, Carlos A. Fuzo, Pedro V. da Silva-Neto, Diana M. Toro, Camila O. S. Souza, Thais F. C. Fraga-Silva, Luiz Gustavo Gardinassi, Jonatan C. S. de Carvalho, Nicola T. Neto, Ingryd Carmona-Garcia, Camilla N. S. Oliveira, Cristiane M. Milanezi, Viviani Nardini Takahashi, Thais Canassa De Leo, Lilian C. Rodrigues, Cassia F. S. L. Dias, Ana C. Xavier, Giovanna S. Porcel, Isabelle C. Guarneri, Kamila Zaparoli, Caroline T. Garbato, Jamille G. M. Argolo, Ângelo A. F. Júnior, Marley R. Feitosa, Rogerio S. Parra, José J. R. da Rocha, Omar Feres, Fernando C. Vilar, Gilberto G. Gaspar, Rafael C. da Silva, Leticia F. Constant, Fátima M. Ostini, Alessandro P. de Amorim, Augusto M. Degiovani, Dayane P. da Silva, Debora C. Nepomuceno, Rita C. C. Barbieri, Isabel K. F. M. Santos, Sandra R. C. Maruyama, Elisa M. S. Russo, Angelina L. Viana, Ana P. M. Fernandes, Vânia L. D. Bonato, Cristina R. B. Cardoso, Carlos A. Sorgi, Marcelo Dias-Baruffi, Lúcia H. Faccioli

## Abstract

Cytokine storms and hyperinflammation, potentially controlled by glucocorticoids, occur in COVID-19; the roles of lipid mediators and acetylcholine (ACh) and how glucocorticoid therapy affects their release in Covid-19 remain unclear. Blood and bronchoalveolar lavage (BAL) samples from SARS-CoV-2- and non-SARS-CoV-2-infected subjects were collected for metabolomic/lipidomic, cytokines, soluble CD14 (sCD14), and ACh, and CD14 and CD36-expressing monocyte/macrophage subpopulation analyses. Transcriptome reanalysis of pulmonary biopsies was performed by assessing coexpression, differential expression, and biological networks. Correlations of lipid mediators, sCD14, and ACh with glucocorticoid treatment were evaluated. This study enrolled 190 participants with Covid-19 at different disease stages, 13 hospitalized non-Covid-19 patients, and 39 healthy-participants. SARS-CoV-2 infection increased blood levels of arachidonic acid (AA), 5-HETE, 11-HETE, sCD14, and ACh but decreased monocyte CD14 and CD36 expression. 5-HETE, 11-HETE, cytokines, ACh, and neutrophils were higher in BAL than in circulation (fold-change for 5-HETE 389.0; 11-HETE 13.6; ACh 18.7, neutrophil 177.5, respectively). Only AA was higher in circulation than in BAL samples (fold-change 7.7). Results were considered significant at P<0.05, 95%CI. Transcriptome data revealed a unique gene expression profile associated with AA, 5-HETE, 11-HETE, ACh, and their receptors in Covid-19. Glucocorticoid treatment in severe/critical cases lowered ACh without impacting disease outcome. We first report that pulmonary inflammation and the worst outcomes in Covid-19 are associated with high levels of ACh and lipid mediators. Glucocorticoid therapy only reduced ACh, and we suggest that treatment may be started early, in combination with AA metabolism inhibitors, to better benefit severe/critical patients.

## Introduction

Individuals Covid-19 may present asymptomatically or with manifestations ranging from acute respiratory distress syndrome to systemic hyperinflammation and organ failure, events attributed to cytokine storms^1^. Free polyunsaturated fatty acids, such as AA and derivative eicosanoids, regulate inflammation^2, 3^, yet their role in Covid-19 has not been well investigated.

ACh, which is released by nerves^4^, leukocytes^5^, and airway epithelial cells^6^, regulates metabolism^7^, cardiac function^8^, airway inflammation^9^, and cytokine production^10^, all of which occur in Covid-19. It is known that eicosanoids stimulate ACh release^3^, but crosstalk between cholinergic and lipid mediator pathways in Covid-19 still need to be clarified.

In this study, levels of lipid mediators, ACh, and other inflammatory markers in blood and BAL from patients with Covid-19 who were treated or not with glucocorticoids were compared to those of non-Covid-19 and healthy-participants. Moreover, lung biopsy transcriptome reanalysis data from Covid-19 and non-Covid-19 patients corroborated our findings.

## Methods

### Study design and blood collection

This observational, analytic, and transversal study was conducted from June to November 2020. All participants were over 16 years old and chosen according to the inclusion and exclusion criteria described in Table S1 and in the protocol, after providing signed consent. Blood samples collected from patients positive for Covid-19 (n=190) were analyzed by RT-qPCR (Biomol OneStep/Covid-19 kit; Institute of Molecular Biology of Paraná - IBMP Curitiba/PR, Brazil) using nasopharyngeal swabs and/or serological assays to detect IgM/IgG/IgA (SARS-CoV-2® antibody test; Guangzhou Wondfo Biotech, China). Samples obtained from a cohort of SARS-CoV-2-negative healthy participants were used as controls (n=39). Participants positive for Covid-19 were categorised as asymptomatic-mild (n=43), moderate (n=44), severe (n=54), or critical (n=49). The criteria for the clinical classification of patients were defined at the time of sample collection, as shown in Table S1. Peripheral blood samples were obtained by venous puncture from patients upon their first admission and/or during the period of hospitalisation at two medical centres, *Santa Casa de Misericordia de Ribeirão Preto* and *Hospital Sao Paulo* at Ribeirão Preto, São Paulo State, Brazil. Blood samples from healthy controls and asymptomatic-mild non-hospitalized participants were collected either at the Centre of Scientific and Technological Development “Supera Park” (Ribeirão Preto, São Paulo State, Brazil) or in the home of patients receiving at-home care. The plasma was separated from whole blood samples and stored at −80°C. For lipidomic and metabolomic analyses, 250 µL of plasma was stored immediately in methanol (1:1 *v/v*). Lipidomic and metabolomic analyses were performed by mass spectrometry (LC-MS/MS), while a cytokines, sCD14, and ACh were quantified using CBA flex Kit (flow cytometer assay) or commercial ELISA. The expression levels of CD14, CD36, CD16, and HLA-DR in cells were evaluated by flow cytometry following the gate strategy (Figure S2).

### Ethical considerations

All participants provided written consent in accordance with the regulations of the *Conselho Nacional de Pesquisa em Humanos* (CONEP) and the Human Ethical Committee from *Faculdade de Ciências Farmacêuticas de Ribeirão Preto* (CEP-FCFRP-USP). The research protocol was approved and received the certificate of Presentation and Ethical Appreciation (CAAE: 30525920.7.0000.5403). The sample size was determined by the convenience of sampling, availability at partner hospitals, agreement to participate, and the pandemic conditions within the local community (more information in the Protocol).

### Bronchoalveolar lavage fluid (BAL) collection and processing

BAL fluids were collected from hospitalised Covid-19 patients at the severe or critical stages of disease (n=32) to assess their lung immune responses. Control samples were obtained from hospitalised intubated donors negative for SARS-CoV-2 (n=13) (as certified by SARS-CoV-2-negative PCR), referred to as non-Covid-19 patients, who were intubated because of the following primary conditions: bacterial pneumonia, abdominal septic shock associated with respiratory distress syndrome, pulmonary atelectasis due to phrenic nerve damage, or pulmonary tuberculosis. BAL fluid was collected as previously described^11^, using a siliconized polyvinylchloride catheter (Mark Med, Porto Alegre, Brazil) with a closed Trach Care endotracheal suction system (Bioteque Corporation, Chirurgic Fernandes Ltd., Santana Parnaíba, Brazil) and sterile 120 mL polypropylene flask (Biomeg-Biotec Hospital Products Ltd., Mairiporã, Brazil) under aseptic conditions. Approximately 5–10 mL of bronchoalveolar fluid was obtained and placed on ice for processing within 4 h. The BAL fluids were placed into 15-mL polypropylene collection tubes and received half volume of their volume of phosphate buffered saline (PBS) 0.1 M (2:1 *v/v*) in relation to the total volume of each sample. After centrifugation (700 × *g*, 10 min), the supernatants of the BAL fluid were recovered and stored at –80°C. For lipidomic and metabolomic analyses, 250 µL of these supernatants were stored immediately in methanol (1:1 *v/v*). Subsequently, the remaining BAL fluid was diluted in 10 mL of PBS and gently filtered through a 100-µm cell strainer (Costar, Corning, NY, USA) using a syringe plunger. The resulting material was used for cytokine and acetylcholine (ACh) quantification. The BAL fluids were centrifuged (700 × *g*, 10 min) and the red blood cells were lysed using 1 mL of ammonium chloride (NH4Cl) buffer 0.16 M for 5 min. The remaining airway cells were washed with 10 mL of PBS, resuspended in PBS–2% heat-inactivated foetal calf serum, and counted with Trypan blue using an automated cell counter (Countess, Thermo Fisher Scientific, Waltham, MA, USA). The leukocyte numbers were adjusted to 1 × 10^9^ cells/L for differential counts and 1 × 10^6^ cells/mL for flow cytometry analysis. All procedures were performed in a Level 3 Biosafety Facility (*Departamento de Bioquímica e Imunologia, Faculdade de Medicina de Ribeirão Preto, Universidade de São Paulo*).

### Data collection

The electronic medical records of each patient were carefully reviewed. Data included sociodemographic information, comorbidities, medical history, clinical symptoms, routine laboratory tests, immunological tests, chest computed tomography (CT) scans, clinical interventions, and outcomes (more information in the Protocol). The information was documented on a standardised record form, as indicated in Tables S1, S2, and S3. Data collection of laboratory results included first-time examinations within 24 h of admission, defined as the primary endpoint. The secondary endpoint was clinical outcome (death or recovery).

### Clinical laboratory collection

For hospitalised patients, blood examinations were performed by clinical analysis laboratories at their respective hospitals. Blood examinations of healthy participants and non-hospitalized patients were performed at *Serviço de Análises Clínicas* (SAC), *Departamento de Análises Clínicas, Toxicológicas e Bromatológicas* of the *Faculdade de Ciências Farmacêuticas de Ribeirão Preto, Universidade de São Paulo, Ribeirão Preto, São Paulo,* Brazil. The blood samples were used to measure for liver and kidney function, myocardial enzyme spectrum, coagulation factors, red blood cells, haemoglobin, platelets, and total and differential leukocytes using automated equipment. Similarly, the absolute numbers of leukocytes in the BAL fluid were determined in a Neubauer Chamber with Turkey solution. For the counts of differential leukocytes in the BAL, 100 µL of the fluid was added to cytospin immediately after collection to avoid any interference on cell morphology. Differential leukocyte counts were conducted using an average of 200 cells after staining with Fast Panoptic (LABORCLIN; Laboratory Products Ltd, Pinhais, Brazil) and examined under an optical microscope (Zeiss EM109; Carl Zeiss AG, Oberkochen, Germany) with a 100× objective (immersion oil) equipped with a Veleta CCD digital camera (Olympus Soft Imaging Solutions Gmbh, Germany) and ImageJ (1.45s) (National Institutes of Health, Rockville, MD, USA)^12^. Lymphocytes, neutrophils, eosinophils, and monocytes/macrophages were identified and morphologically characterised, and their lengths and widths were measured (100×).

### High-performance liquid chromatography coupled with tandem Mass Spectrometry (LC-MS/MS) assay

#### Reagents

Eicosanoids, free fatty acids (AA, EPA, and DHA), and metabolites as molecular weight standards (MWS) and deuterated internal standards were purchased from Cayman Chemical Co. (Ann Arbor, MI, USA). HPLC-grade acetonitrile (ACN), methanol (MeOH), and isopropanol were purchased from Merck (Kenilworth, NJ, USA). Ultrapure deionised water (H2O) was obtained using a Milli-Q water purification system (Merck-Millipore, Kenilworth, NJ, USA). Acetic acid (CH3COOH) and ammonium hydroxide (NH4OH) were obtained from Sigma Aldrich (St. Louis, MO, USA).

#### Sample preparation and extraction

The plasma (250 μL) in EDTA-containing tubes (Vacutainer® EDTA K2; BD Diagnostics, Franklin Lakes, NJ, USA) and BAL (250 μL) samples were stored in MeOH (1:1, *v/v*) at ‒80°C. Three additional volumes of ice-cold absolute MeOH were added to each sample overnight at ‒20°C for protein denaturation and after lipid solid-phase extraction (SPE). To each sample, 10 μL of internal standard (IS) solution was added, centrifuged at 800 × *g* for 10 min at 4°C. The resulting supernatants were collected and diluted with deionised water (ultrapure water; Merck-Millipore, Kenilworth, NJ, USA) to obtain a MeOH concentration of 10% (*v/v*). In the SPE extractions, a Hypersep C18-500 mg column (3 mL) (Thermo Scientific-Bellefonte, PA, USA) equipped with an extraction manifold collector (Waters-Milford, MA, USA) was used. The diluted samples were loaded into the pre-equilibrated column and washing using 2 mL of MeOH and H2O containing 0.1% acetic acid, respectively. Then, the cartridges were flushed with 4 mL of H2O containing 0.1% acetic acid to remove hydrophilic impurities. The lipids that had been adsorbed on the SPE sorbent were eluted with 1 mL of MeOH containing 0.1% acetic acid. The eluates solvent was removed in vacuum (Concentrator Plus, Eppendorf, Germany) at room temperature and reconstituted in 50 μL of MeOH/H2O (7:3, *v/v*) for LC-MS/MS analysis.

#### LC-MS/MS analysis and lipids data processing

Liquid chromatography was performed using an Ascentis Express C18 column (Supelco, St. Louis, MO, USA) with 100 × 4.6 mm and a particle size of 2.7 μm in a high-performance liquid chromatography (HPLC) system (Nexera X2; Shimadzu, Kyoto, Japan). Then, 20 μL of extracted sample was injected into the HPLC column. Elution was carried out under a binary gradient system consisting of Phase A, comprised of H2O,ACN, and acetic acid (69.98:30:0.02, *v/v/v*) at pH 5.8 (adjusted with NH4OH), and Phase B, comprised of ACN and isopropanol (70:30, *v/v*). Gradient elution was performed for 25 min at a flow rate of 0.5 mL/min. The gradient conditions were as follows: 0 to 2 min, 0% B; 2 to 5 min, 15% B; 5 to 8 min, 20% B; 8 to 11 min, 35% B; 11 to 15 min, 70% B; and 15 to 19 min, 100% B. At 19 min, the gradient was returned to the initial condition of 0% B, and the column was re-equilibrated until 25 min. During analysis, the column samples were maintained at 25°C and 4°C in the auto-sampler. The HPLC system was directly connected to a TripleTOF 5600+ mass spectrometer (SCIEX-Foster, CA, USA). An electrospray ionisation source (ESI) in negative ion mode was used for high-resolution multiple-reaction monitoring (MRM^HR^) scanning. An atmospheric-pressure chemical ionisation probe (APCI) was used for external calibrations of the calibrated delivery system (CDS). Automatic mass calibration (<2 ppm) was performed periodically after each of the five sample injections using APCI Negative Calibration Solution (Sciex-Foster, CA, USA) injected via direct infusion at a flow rate of 300 μL/min. Additional instrumental parameters were as follows: nebuliser gas (GS1), 50 psi; turbo gas (GS2), 50 psi; curtain gas (CUR), 25 psi; electrospray voltage (ISVF), ‒4.0 kV; temperature of the turbo ion spray source, 550°C. The dwell time was 10 ms, and a mass resolution of 35,000 was achieved at *m/z* 400. Data acquisition was performed using Analyst^TM^ software (SCIEX-Foster, CA, USA). Qualitative identification of the lipid species was performed using PeakView^TM^ (SCIEX-Foster, CA, USA). MultiQuant^TM^ (SCIEX-Foster, CA, USA) was used for the quantitative analysis, which allows the normalisation of the peak intensities of individual molecular ions using an internal standard for each class of lipid. The quantification of each compound was performed using internal standards and calibration curves, and the specific mass transitions of each lipid were determined according to our previously published method^13^. The final concentration of lipids was normalised by the initial volume of plasma or BAL fluid (ng/mL).

#### Metabolomics analysis

Metabolite was extracted and samples were transferred to autosampler vials for LC–MS analysis using TripleTOF5600+ Mass Spectrometer (Sciex-Foster, CA, USA) coupled to an ultra-high-performance liquid chromatography (UHPLC) system (Nexera X2; Shimadzu, Kyoto, Japan). Reverse-phase chromatography was performed similarly to lipids analyses above. Mass spectral data were acquired with negative electrospray ionisation, and the full scan of mass-to-charge ratio (*m/z*) ranged from 100 to 1500. Proteowizard software^14^ was used to convert the wiff files into *mz* XML files. Peak peaking, noise filtering, retention time, *m/z* alignment, and feature quantification were performed using apLCMS^15^. Three parameters were used to define a metabolite feature: mass-to-charge ratio (*m/z*), retention time (min), and intensity values. Data were log2 transformed and only features detected in at least 50% of samples from one group were used in further analyses. Missing values were imputed using half the mean of the feature across all samples. Mummichog (version 2) was used for metabolic pathway enrichment analysis (mass accuracy under 10 ppm)^16^.

### Acetylcholine measurement

ACh was measured in heparinized plasma (SST® Gel Advance®; BD Diagnostics, Franklin Lakes, NJ, USA) and in BAL using a commercially available immunofluorescence kit (ab65345; Abcam, Cambridge, UK) according to the manufacturer’s instructions. Briefly, ACh was converted to choline by adding the enzyme acetylcholinesterase to the reaction, which allows for total and free-choline measurement.

The amount of ACh present in the samples was calculated by subtracting the free choline from the total choline. The products formed in the assay react with the choline probe and can be measured by fluorescence with excitation and emission wavelengths of 535 and 587 nm, respectively (Paradigm Plate Reader; SpectraMax, San Diego, CA, USA). The concentration of ACh was analysed using SoftMax® software (SpectraMax, Molecular Devices, Sunnyvale, CA, USA), expressed as pmol.mL^-1^.

### Soluble CD14 (sCD14) measurement

Samples from heparinized plasma (SST® Gel Advance®; BD Biosciences, Franklin Lakes, NJ, USA) were placed in 96-well plates. The concentration of sCD14 was determined using an ELISA kit (DY383; R&D Systems, Minneapolis, MN, USA), following the manufacturer’s instructions, expressed as pg.mL^-1^.

### Flow Cytometry

Uncoagulated blood samples in EDTA-containing tubes (Vacutainer® EDTA K2; BD Biosciences) were processed for flow cytometry analysis of circulating leukocytes. Whole blood (1 mL) was separated and red blood cells were lysed using RBC lysis buffer (Roche Diagnostics GmbH, Mannheim, GR). Leukocytes were washed in PBS containing 5% foetal bovine serum (FBS) (Gibco™, USA), centrifuged, and resuspended in Hank’s balanced salt solution (Sigma-Aldrich, Merck, Darmstadt, Germany) containing 5% FBS, followed by surface antigen staining. Similarly, cells obtained from BAL fluid were processed for flow cytometry assays. Briefly, cells were stained with Fixable Viability Stain 620 (1:1000) (BD Biosciences) and incubated with monoclonal antibodies specific for CD14 (1:100) (M5E2; Biolegend), HLA-DR (1:100) (G46-6; BD Biosciences), CD16 (1:100) (3G8; Biolegend), and CD36 (1:100) (CB38, BD Biosciences) for 30 min at 4°C. Stained cells were washed and fixed with BD Cytofix™ Fixation Buffer (554655; BD Biosciences, San Diego, CA, USA). Data acquisition was performed using a LSR-Fortessa™ flow cytometer (BD Biosciences, San Jose, CA, USA) and FACS-Diva software (version 8.0.1) (BD Biosciences, Franklin Lakes, NJ, USA). For the analysis, 300,000 events were acquired for each sample. Data were evaluated using FlowJo® software (version 10.7.0) (Tree Star, Ashland, OR, USA) to calculate the cell frequency, dimensionality reduction, and visualisation using t-distributed stochastic neighbour embedding. Gate strategy performed as described before ^17^, as shown in Figure S2.

### Cytokine Measurements

The cytokines interleukin (IL)-6, IL-8, IL-1ß, IL-10, and tumour necrosis factor (TNF) were quantified in heparinized plasma and BAL fluid samples using a BD Cytometric Bead Array (CBA) Human Inflammatory Kit (BD Biosciences, San Jose, CA, USA), according to manufacturer’s instructions. Briefly, after sample processing, the cytokine beads were counted using a flow cytometer (FACS Canto TM II; BD Biosciences, San Diego, CA, USA), and analyses were performed using FCAP Array (3.0) software (BD Biosciences, San Jose, CA, USA). The concentrations of cytokines were expressed as pg.mL^-1^.

### Re-analysis of transcriptome data from lung biopsies of patients with Covid-19

To gain a better understanding of the correlation between the altered concentrations of ACh, AA, and AA-metabolites detected in the plasma and BAL fluid of severe/critical Covid-19 patients, we performed a new analysis by re-using a previously published transcriptome open dataset ^18^, deposited in the Gene Expression Omnibus repository under accession no. GSE150316 ^19^. We used transcriptome data from lung samples (n=46) from patients with Covid-19 (n=15), seven of which displayed a low viral load and eight a high viral load, and non-Covid-19 patients (n=5) with other pulmonary illnesses (negative control). Patients with a high viral load had meantime periods of hospital stays (3.6 ± 2.2 days) and duration of illness (7.2 ± 3.02 days) shorter than the patients with low viral load (14 ± 7.9 and 19 ± 4.9 days, respectively), as described by the authors of the public data source ^18^. Hence, for analysis purposes, all patient samples were grouped into four classifications: Covid-19 (CV), Covid-19 low viral load (CVL), Covid-19 high viral load (CVH), and non-Covid-19 (NCV). The strategy for reanalysing the transcriptome was implemented according to three consecutive steps: (i) co-expression analysis, (ii) differential expression analysis, and (iii) biological network construction. Initially, for the co-expression study, normalised transcriptome data in log2 of reads per million (RPM) were filtered by excluding non-zero counts in at least 20% of the samples. Next, the selected genes were explored in the R package Co-Expression Modules identification Tool (CEMITool) ^20^, using a *p*-value of 0.05 as the threshold for filtering. Then, the co-expression modules were analysed for the occurrence of ACh and AA genes list obtained from the Reactome pathways ^21^, as well as the Covid-19-related genes obtained from the literature (Supplementary Appendix I). Next, differential gene expression between samples from the lung biopsy transcriptome (CV, NCV, CVL, and CVH) was measured using the DESeq2 package ^22^, with *p*-values adjusted using the Benjamini and Hochberg method ^23^. The list of differentially express genes (DEGs) generated for all comparisons was filtered from the genes listed in Supplementary Appendix I, considering the values of log2 of fold-change (FC) greater than 1 (|log2(FC)|>1) and adjusted *p*<0.05. Finally, a first-order biological network was constructed using co-expression module(s) containing genes associated with the ACh and AA pathways to characterise the interplay between these mediators in combination with Covid-19 severity markers, as well as to identify relevant DEGs and hub genes in this network, using the BioGRID repository ^24^. The networks were constructed, analysed, and graphically represented using the R packages igraph ^25^, Intergraph ^26^, and ggnetwork ^27^.

Due to the substantial influence of glucocorticoid treatment on the levels of some mediators, we measured the sensitivity of genes from the differential expression analysis between CV samples from patients who underwent treatment (CTC, three patients and ten samples) and patients who were not treated (NCTC, 12 patients and 36 samples), as previously described ^18^.

### Statistical Analysis

Two-tailed tests were used for the statistical analysis, with a significance value of *p* <0.05 and a confidence interval of 95%. The data were evaluated for a normal distribution using the Kolmogorov–Smirnov test. The parametric data were analysed using unpaired t-tests (for two groups) or one-way ANOVA followed by Tukey’s multiple comparison tests for three or more groups simultaneously. For data that did not display a Gaussian distribution, Mann-Whitney (for two-group comparisons) or Kruskal-Wallis testes were used, followed by Dunn’s post-tests for analysis among three or more groups. The cytokine network data in patients with Covid-19 were analysed using significant Spearman’s correlations at *p*<0.05. Data were represented by connecting edges to highlight positive strong (*r* ≥ 0.68; thick continuous line), moderate (0.36 ≥ *r* < 0.68; thinner continuous line), or weak (0 > *r* < 0.36; thin continuous line) and negative strong (*r* ≤ ‒0.68; thick dashed line), moderate (‒0.68 > *r* ≤ ‒0.36; thinner dashed line), or weak (-0.36 < *r* > 0; thin dashed line), as proposed previously ^28, 29^. The absence of a line indicates the non-existence of the relationship. The Venn diagrams were elaborated using the online tool Draw Venn Diagram (http://bioinformatics.psb.ugent.be/webtools/Venn/).

The results were tabulated using GraphPad Prism software (version 8.0) and the differences were considered statistically significant at *p*<0.05. See the Additional Statistical Report section for more information. Some of the confounding variables associated with Covid-19 (age, sex, obesity, hypertension, and diabetes mellitus) were analysed for their potential impacts on the main analytical procedures of this study, such as ACh, AA, 5-HETE, and 11-HETE measurements of the plasma (healthy, asymptomatic-to-mild, moderate, severe, and critical patients) and BAL (severe and critical patients) samples. This analysis was performed using the Kruskal-Wallis, Mann-Whitney, Spearman’s correlation, or Chi-square (χ2) tests (Table S7-S9).

## Results

### Study Population

This study enrolled 39 healthy-participants, 13 hospitalized non-Covid-19, and 190 Covid-19 patients aged 16-96 years from April to November 2020. The 190 Covid-19 patients were categorized as having asymptomatic-to-mild (n=43), moderate (n=44), severe (n=54), or critical (n=49) disease (Table S2).

### Covid-19 Modifies Circulating Soluble Mediators and Cell Populations

To determine whether SARS-CoV-2 infection alters the metabolism of lipid mediators, we used high-resolution sensitive mass spectrometry to perform targeted eicosanoid analysis and nontargeted metabolomics using plasma from healthy-participants and Covid-19 patients. In total, 8,791 metabolite features were present in at least 50% of all samples, and the relative abundance of 595 metabolite features (FDR adjusted P<0.05) was altered in the groups studied (Figure 1A). Two-way hierarchical clustering based on these significant metabolite features resulted in three clear clusters: one for severe/critical Covid-19 patients, one for healthy-participants and one for asymptomatic-to-mild and moderate Covid-19 (Figure S1A). Pathway analysis revealed the top significant metabolic pathways to be enriched in features involved in fatty acid biosynthesis, metabolism, activation, and oxidation (Figure 1B). Compared to healthy-participants, tentative metabolite annotations suggested an increased abundance of fatty acids (FFAs), such as linoleic acid, tetradecanoate, dodecanoate and AA, in COVID-19 (Figure 4C-F). Among the identified lipids, AA was the most abundant, and its levels correlated with the severity of Covid-19 (Figure 1F). Linoleic acid can be metabolized to AA, which in turn is a substrate for eicosanoids, such as 5-hydroxyeicosatetraenoic acid (5-HETE) and 11-hydroxyeicosatetraenoic acid (11-HETE) (Figure 1G, 1H); both molecules with function in the immune response. Overall, the increased plasma levels of AA in Covid-19 indicate that it predicts disease severity.

**Figure 1.**
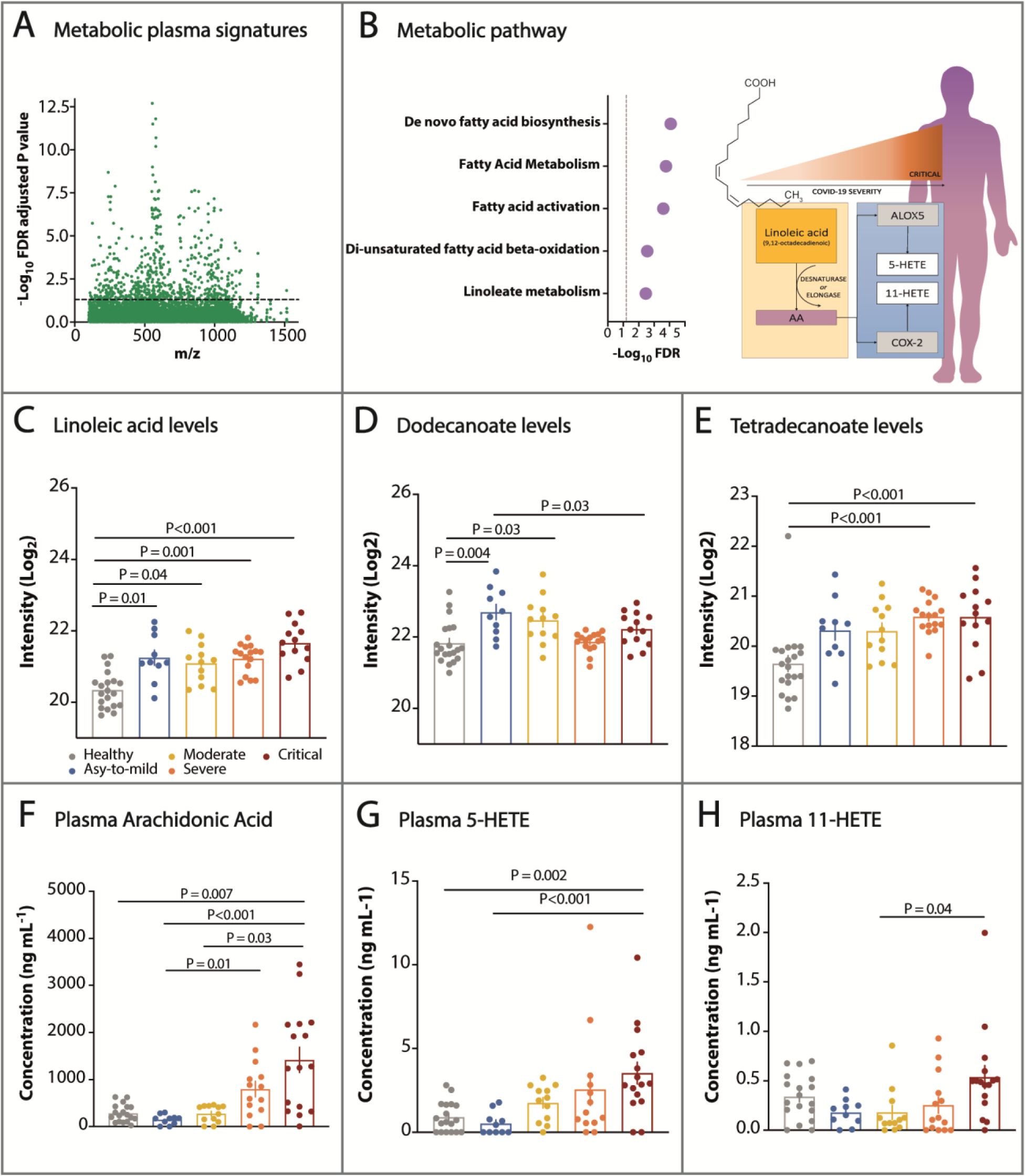
Metabolomic and lipidomic analysis revealed increased levels of circulating fatty acids, arachidonic acid (AA), 5-HETE and 11-HETE in patients with Covid-19. Plasma samples were collected from healthy-participants (n=20) and patients with asymptomatic-to-mild (n=10), moderate (n=12), severe (n=16), or critical (n=13) disease. (A) A Manhattan plot for the differential abundance of metabolite features in different groups of patients with Covid-19 and healthy participants (false discovery rate (FDR) of 595, adjusted P<0.05, above the dashed line) and (B, left panel) metabolic pathway enrichment of significant metabolite features based on data from untargeted mass spectrometry. Differential abundance was calculated using the *limma* package for R, and FDR was controlled using the Benjamini-Hochberg method. Mummichog software v2.3.3 was used for pathway enrichment analysis. (B, right panel) Schematic illustration of the metabolic pathways involved in the production of hydroxyeicosatetraenoic acids (HETEs), such as 5-HETE and 11-HETE, from arachidonic (AA) and linoleic acid metabolism, which discriminate the severity of Covid-19. (C-E) Plasma metabolomics indicating an increase in free fatty acids in Covid-19. Plasma obtained from healthy participants (n=18) and patients with asymptomatic-to-mild (n=10), moderate (n=12), severe (n=14), or critical (n=16) disease (F-H) was subjected to lipidomics analysis using targeted mass spectrometry, confirming the elevation of AA, 5-HETE and 11-HETE, according to the severity of Covid-19. Data are expressed as the mean ± SEM. Differences in (C-H) were considered significant at P<0.05 according to Kruskal–Wallis analysis followed by Dunn’s posttest, and specific P-values are shown in each figure.

As eicosanoids induce cell recruitment and regulate immune responses, we next determined the profile of immune cells and soluble mediators in whole blood and plasma of patients with Covid-19 and healthy-participants. According to whole-blood analysis, absolute leukocyte and neutrophil counts (Figure 2A, 2B) were significantly higher but lymphocyte counts (Figure 2C) significantly lower in patients with severe/critical disease than in those with moderate disease. Eosinophil (Figure 2D) and basophil counts (Figure 2E) were reduced severe disease compared to asymptomatic and moderate disease.

**Figure 2.**
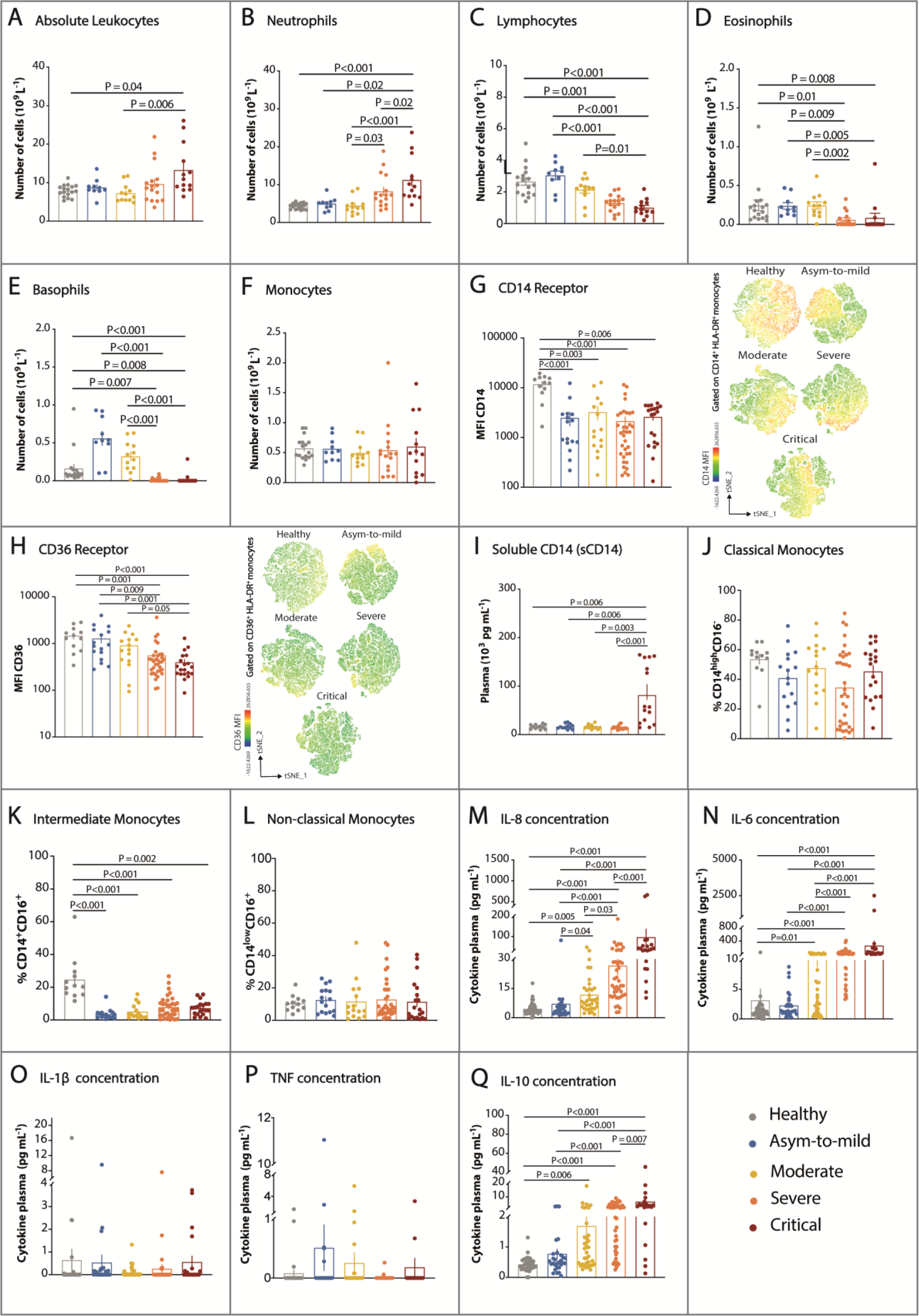
Systemic markers of inflammation determined Covid-19 severity. (A-F) Total and differential leukocyte counts in peripheral blood from healthy-participants (n=17) and from patients with asymptomatic-to-mild (n=10), moderate (n=12), severe (n=16) or critical (n=13) disease showed that Covid-19 modified the numbers of distinct leukocyte populations. (G-H) Flow cytometry analysis of CD14^+^HLA-DR circulating monocytes by t-distributed stochastic neighbour embedding indicated reduced (G) CD14 and (H) CD36 mean fluorescence intensity in asymptomatic-to-mild (n=16), moderate (n=15), severe (n=35), and critical (n=20) Covid-19 patients compared to healthy participants (n=12). (I) Soluble CD14 (sCD14), as determined by ELISA) in healthy participants (n=12), asymptomatic-to-mild (n=12), moderate (n=13), severe (n=15), and critical (n=15) patients, showed increases only in samples from critical Covid-19 patients. (J-L) Classical, intermediate, and nonclassical monocytes determined by flow cytometry analyses of CD14, CD16 and HLA-DR expression in blood cells from healthy participants (n=12), asymptomatic-to-mild (n=16), moderate (n=15), severe (n=35), and critical (n=20) Covid-19 patients revealed a decrease in intermediate monocytes in Covid-19. (M-Q) Cytokines (IL-8, IL-6, IL-1β, TNF and IL-10) quantified in plasma using a Cytometric Bead Array (CBA) of healthy volunteers (n=35), asymptomatic-to-mild (n=29), moderate (n=35), severe (n=42), and critical (n=21) patients demonstrated a distinct cytokine profile according to disease severity. Data are expressed as the mean ± SEM, and differences between groups were calculated using Kruskal–Wallis with Dunn’s multiple comparison post-tests. The specific P-values are displayed in each figure. Differences were considered significant at P<0.05.

Although no differences in total monocyte counts among the groups (Figure 2F) were observed based on CD16, CD14 and HLA-DR, expression of membrane CD14 was reduced in all SARS-CoV-2-infected patients compared to healthy-participants (Figure 2G; Figure S2, S3). In parallel, CD36 expression was decreased in monocytes from patients with severe/critical disease (Figure 2H), but sCD14 was increased only in plasma from critical patients (Figure 2I). We did not detect differences in classic or non-classic monocytes, but the percentage of intermediate monocytes was decreased in all SARS-CoV-2-infected participants compared to healthy-participants (Figure 2J, 2K, 2L). Based on plasma cytokine level analysis, patients with moderate, severe, and critical disease share a Covid-19 cytokine profile defined by increased IL-8, IL-6, and IL-10 (Figure 2M, 2N, 2Q) levels, with unaltered IL-1β and TNF levels (Figure 2O, 2P).

### Covid-19 Induces Strong Lung Responses

We performed measurements of BAL from hospitalized Covid-19 and non-Covid-19 patients. Changes in lung metabolomics induced by SARS-CoV-2 infection (Figure 3A; Figure S1B) included alterations in sphingolipids, beta oxidation of trihydroxyprostanoil-CoA, biosynthesis and metabolism of steroidal hormones, vitamin D3, and glycerophospholipids (Figure 3B). We also evaluated AA and its metabolites. Despite no differences in AA, levels of 5-HETE and 11-HETE in BAL were significantly higher in Covid-19 than in non-Covid-19 patients, though other metabolites did not differ between these groups (Figure 3C). When assessing leukocytes in BAL between the patient groups, we found no differences in total or differential counts, with the exception of lymphocyte numbers (Figure 3D, 3E). In contrast to eicosanoids, cytokine profiles in Covid-19 and non-Covid-19 patients were similar (Figure 3F), suggesting that lipid mediators contribute to the pathophysiological processes induced by SARS-CoV-2. Interestingly, we observed a significant reduction in classical (Figure 3G) and intermediate (Figure 3H) monocytes in BAL from Covid-19 patients. In parallel, CD14 and CD36 expression in monocyte was lower in BAL of Covid-19 than in that of non-Covid-19 patients (Figure 3I, 3J).

**Figure 3.**
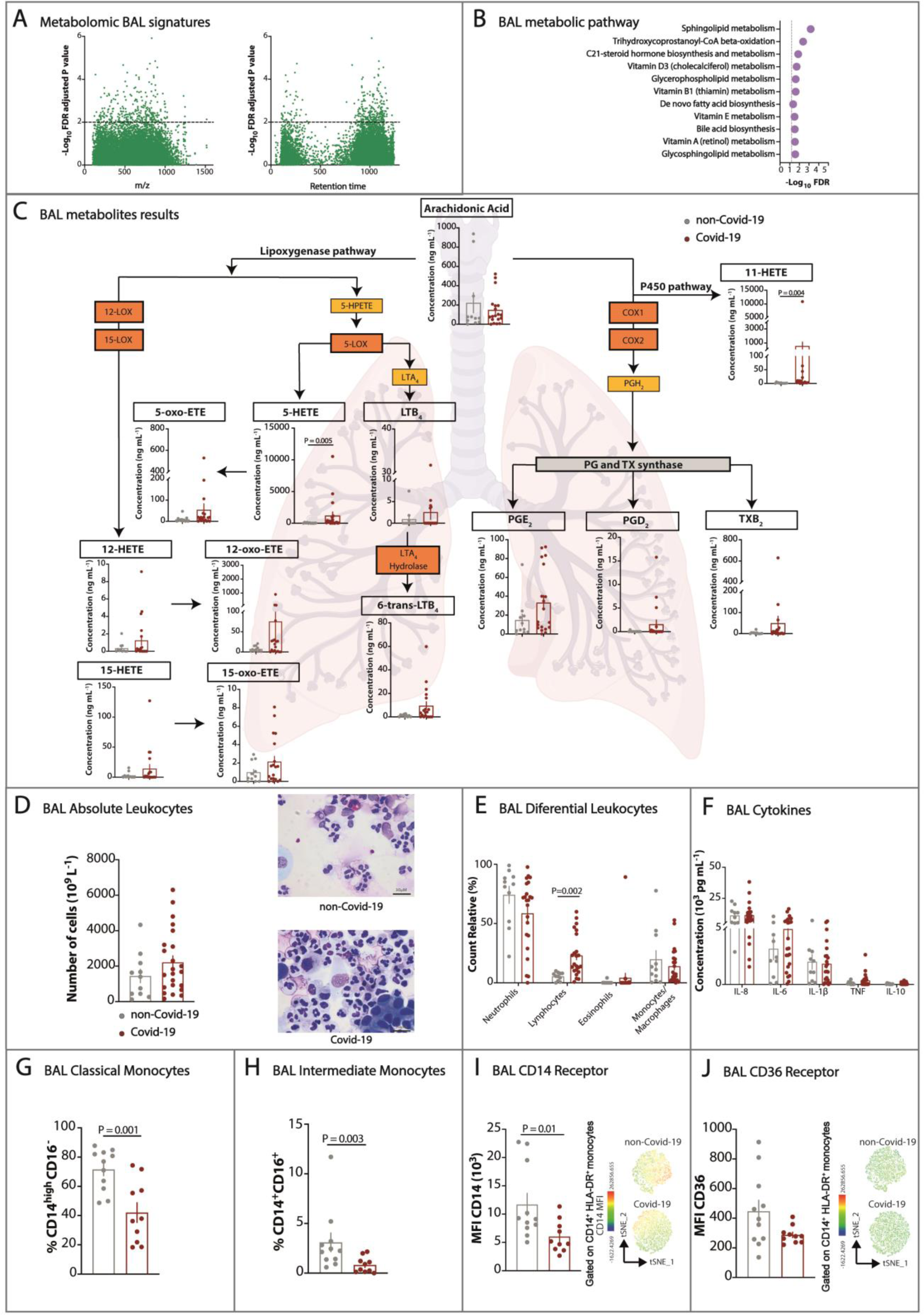
Altered production of lipid mediators and cellular infiltrates in the lungs drove the local response to SARS-CoV-2 infection. BAL was collected from intubated patients with severe/critical confirmed Covid-19 diagnosis and from intubated patients without SARS-CoV-2 infection, which are labelled as Covid-19 and non-Covid-19, respectively. Data from untargeted mass spectrometry demonstrated (A) differential abundance of metabolite features comparing Covid-19 and non-Covid-19 participants (Manhattan plot, 595 at FDR adjusted P<0.05, above the dashed line) and (B) metabolic pathway enrichment of significant metabolite features. Differential abundance was calculated using the *limma* package for R, and the false discovery rate was controlled using the Benjamini-Hochberg method; Covid-19 (n=26) and non-Covid-19 (n=12). Mummichog software v2.3.3 was used for lipid pathway enrichment analysis. (C) Lipid mediators derived from arachidonic acid (AA) metabolism by lipoxygenase (LOX) or cyclooxygenase (COX) in both Covid-19 (n=19) and non-Covid-19 (n=11) samples, as determined by target mass spectrometry, showed a significant increase in 5-HETE and 11-HETE in Covid-19. (D) Total leukocytes in BAL fluid (left panel) and a representative image of Romanowsky staining in these cells (right panel) used for (E) identification of specific cellular populations in Covid-19 (n=23) and non-Covid-19 (n=11) patients showed a similar profile of infiltrating cells. (F) Cytokine concentrations in BAL from intubated Covid-19 patients (n=23) and non-Covid-19 participants (n=11) also revealed a similar profile. (G, H) Classical and intermediate monocytes determined by flow cytometry analyses of CD14, CD16 and HLA-DR expression in BAL cells from Covid-19 (n=10) and non-Covid-19 (n=11) patients demonstrate that both populations are decreased in Covid-19. (I) CD14 and (J) CD36 mean fluorescence intensity (MFI) of CD14^+^HLA-DR gated BAL monocytes is decreased in Covid-19 (n=10) compared to non-Covid-19 (n=11) patients. 5-HPETE: 5-hydroperoxyeicosatetraenoic acid, LTA4: leukotriene A4, LTA4 hydrolase: leukotriene A4 hydrolase, LTB4: leukotriene B4, 6-trans LTB4: 6-trans leukotriene B4, 5-HETE: 5-hydroxyeicosatetraenoic acid, 11-HETE: 11-hydroxyeicosatetraenoic acid, 12-HETE: 12-hydroxyeicosatetraenoic acid, 15-HETE: 15-hydroxyeicosatetraenoic acid, 5-oxo-HETE: 5-oxoeicosatetraenoic acid, 12-oxo-ETE: 12-oxoeicosatetraenoic acid, 15-oxo-HETE: 15-oxoeicosatetraenoic acid, PGH2: prostaglandin H2, PGE2: prostaglandin E2, PGD2: prostaglandin D2, TXB2: thromboxane B, TX synthase: thromboxane. Data are expressed as the mean ± SEM. Differences between groups were calculated using the Mann–Whitney test, and specific P-values are shown in the figure. Differences were considered significant at P<0.05.

### Acetylcholine, Cytokines and Eicosanoids are Higher in the Lung Micro-Environment

We compared systemic and lung responses only in patients with severe/critical Covid-19 and found significantly higher levels of cytokines in BAL than in blood (Figure 4A). When comparing lipid mediator levels in either compartment from the same Covid-19 patients, we detected lower AA but higher levels of 5-HETE and 11-HETE in BAL than in blood (Figure 4B, 4C, 4D). Previous results from our group^3^ have shown that eicosanoids contribute to ACh release. Therefore, we next measured ACh in patients with Covid-19 and observed higher plasma levels of ACh in patients with Covid-19; in addition, ACh was elevated in patients with severe/critical disease compared with those with asymptomatic/moderate disease or healthy-participants (Figure 4E). Unexpectedly, in patients with severe/critical Covid-19, ACh levels in BAL were 10-fold higher than those in serum, and patients treated with glucocorticoids showed decreases in ACh in both compartments (Figure 4F, 4G). Interesting, neutrophil counts were higher in BAL, as these cells produce high levels of 5-HETE and IL-1β, both of which are mediators of ACh release^30, 31^ (Figure 4H). Correlation analysis was then performed to evaluate the relationship between eicosanoids, cytokines, sCD14 and ACh in patients with Covid-19. When comparing blood samples from all patients, we detected strong correlations between ACh *versus* IL-1β and moderate correlations between ACh *versus* AA. A substantial number of interactions between AA and its metabolites and between cytokines and eicosanoids were observed (Figure 4I; Figure S4A). Correlations among all parameters were also observed in BAL (Figure 4J; Figure S4B). To evaluate the benefit of glucocorticoid treatments and their relationship with eicosanoids, CD14 and ACh, we analysed the intersections in a Venn diagram of blood and BAL from severe/critical Covid-19. The results showed that treatment of Covid-19 with glucocorticoids did not have a significant influence on eicosanoid or sCD14 release in patients with more severe stages of disease; in critical patients, however, reductions of 44% and 65% in ACh levels in blood and BAL, respectively, were observed (Figure 4K-4Q; Figure S5). These findings suggest that the use of glucocorticoids has a positive effect on resolution of the inflammatory process because they reduce ACh release, which directly or indirectly stimulates cell recruitment and proinflammatory mediator release.

**Figure 4.**
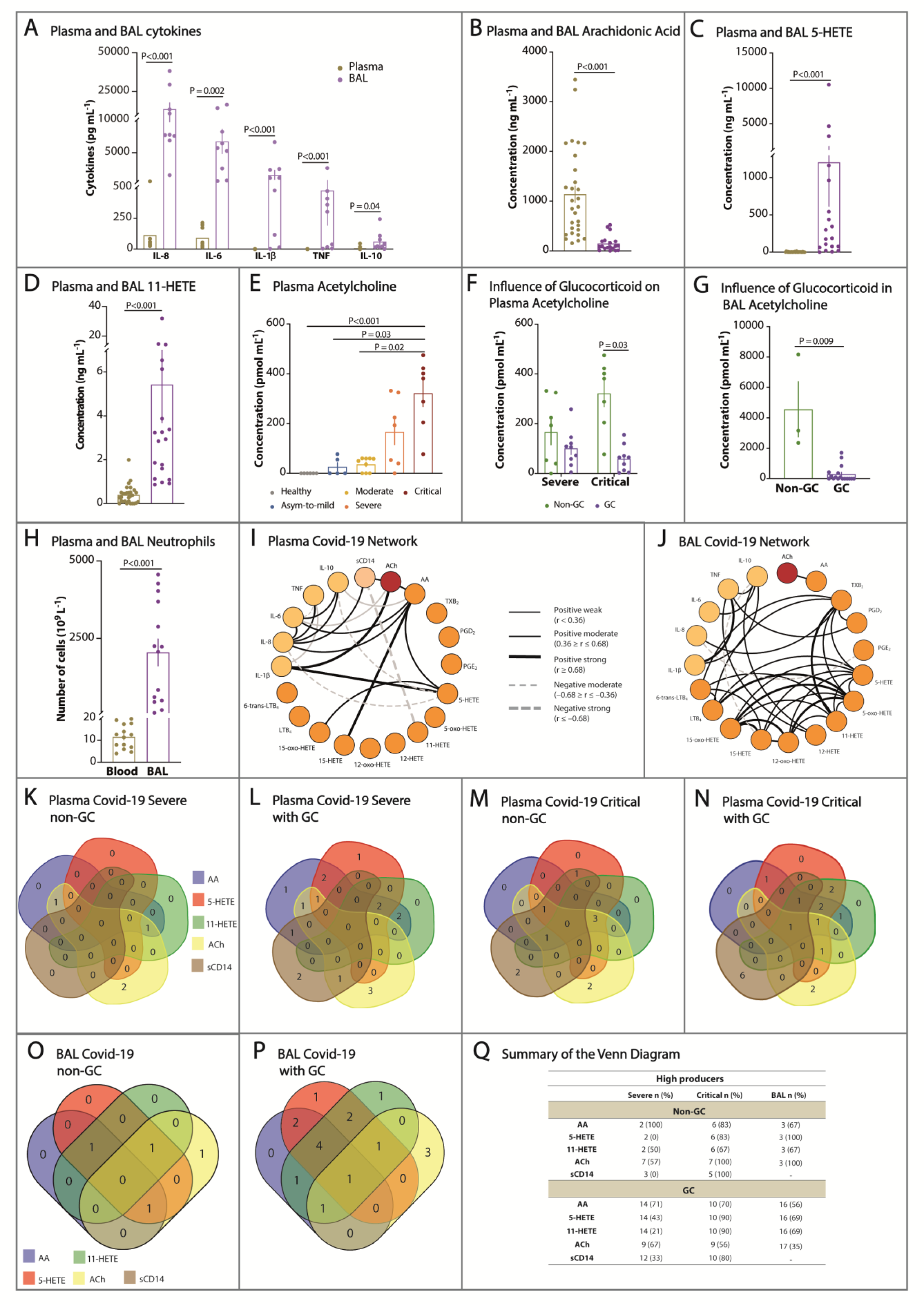
Severe and critical phases of Covid-19 correlated with increased systemic and lung acetylcholine and lipid mediators, but only ACh was diminished by glucocorticoids. Comparison of systemic and local lung responses was analysed using blood and BAL samples from patients with severe/critical Covid-19 and showed that among (A) cytokines (plasma n=9; BAL n=9), (B) AA (plasma n=29; BAL n=19), (C) 5-HETE (plasma n=29; BAL n=19), and (D) 11-HETE (plasma n=29; BAL n=19), levels of only AA were higher in blood. (E) The ACh concentration (pmol.mL^-1^) in heparinized plasma from healthy-participants (n=6) compared to plasma from SARS-CoV-2-infected patients not treated with glucocorticoids (non-GC) classified as having asymptomatic-to-mild (n=5), moderate (n=9), severe (n=7), and critical (n=7) disease showed an increase based on disease severity. (F) ACh in the plasma of patients with Covid-19 at severe/critical stages of the disease and treated (GC, n=18) or not (non-GC, n=14) with glucocorticoids shows decreased release in response to the treatment. (G) Comparison of ACh in BAL from SARS-CoV-2-infected severe/critical patients treated (CG, n=17) or not (non-GC, n=3) with glucocorticoids confirmed inhibition of the neurotransmitter by the treatment. Comparison of neutrophil numbers from blood (n=14) and BAL (n=14) of patients with severe/critical disease demonstrated increased neutrophil infiltration in the lungs. Data are expressed as the mean ± SEM. Differences were considered significant at P<0.05 according to (A) and (E) Kruskal–Wallis tests followed by Dunn’s posttest. (B, C, D, F and G) Student’s *t*-tests, Mann–Whitney, (H) Wilcoxon matched-pairs signed rank test. Concentrations of ACh in the blood of patients with severe and critical disease and not treated with glucocorticoids were the same, as shown in panels (E) and (F). Blood and BAL were collected during hospitalization, on average 6 to 17 days after admission, and glucocorticoids (methylprednisolone; range 40 to 500 mg/kg/day, or dexamethasone; range 1.5 to 6.0 mg/kg/day) were administered intravenously. Interaction networks between various pairs of mediators quantified in (I) blood (n=151) or (J) BAL (n=32) from our Covid-19 cohort, as constructed using the open access software Cytoscape v3.3 (Cytoscape Consortium, San Diego, CA) and Spearman tests and showing significant correlations depicted by different lines (*r* and P values described in section 7.1). (K-P) Venn diagram constructed according to the online tool Draw Venn Diagram (http://bioinformatics.psb.ugent.be/webtools/Venn/) illustrates high-producing patients in severe and critical stages of disease, with (GC) or without (non-GC) treatment. (K-N) Plasma from patients producing high levels of AA, 5-HETE, 11-HETE, ACh and sCD14 in GC (n=49) or non-GC (n=17) groups. (O, P) BAL from patients producing high levels of AA, 5-HETE, 11-HETE, and ACh in GC (n=23) or non-GC (n=4) groups. For analysis, the global median between controls and patients was considered as the cut-off point. (Q) Summary data of the values used for construction of the Venn diagrams. The table shows the absolute number of patients present at each stage of the disease and whether glucocorticoids were used for each molecule analysed. The percentage of individuals present in each experimental condition is highlighted in brackets.

### Altered Expression of Acetylcholine and Arachidonic Acid Pathway Genes in Lung Biopsies From Some Covid-19 Patients

In this study, we reanalysed the lung biopsy transcriptome from Covid-19 patients to evaluate expression of ACh and AA pathway genes (Supplementary Appendix I). These genes were coexpressed only in a single module (M1) that included genes related to the ACh release cycle, AA metabolism, cholinergic and eicosanoid receptors, and biomarkers of Covid-19 severity (Figure 5A; Table S4; Supplementary Appendix II). Furthermore, expression of nearly all genes was upregulated in some deceased Covid-19 patients with long hospital stays and low viral loads (Figure 5B; Figure S6A; Supplementary Appendix III). These differentially expressed genes populated the biological network and are likely under the action of some hubs (Figure 5C; Supplementary Appendix IV), such as oestrogen receptor II (ESR2) and albumin (ALB). We identified a unique proinflammatory gene expression profile in lung biopsies related to cholinergic and eicosanoid receptors (Figure 5H, 5I; Figure S6B, S6C). In combination with the altered levels of ACh, AA, and AA metabolites found in patient from our cohort, the transcriptome data reported herein strengthen the likelihood that these mediators contribute to Covid-19 severity (Figure 5D-5G). Interestingly, lung samples from Covid-19 patients with short hospital stays and high viral loads did not present this unique expression profile of ACh or AA pathway genes (Figure 5H, 5I; Figure S6B, S6C). Some Covid-19 patients treated versus not with glucocorticoid showed transcript expression of monoglyceride lipase (MGLL) and N-acylethanolamine acid amidase (NAAA) and up-regulation of fatty acid amide hydrolase (FAAH), which are involved in the production of AA from endocannabinoid (cases 3, 9, and 11; Figure 5B; Supplementary Appendix III). Besides, we detected other DEGs in biopsy samples from Covid-19 patients (glucocorticoid-treated versus non-treated) associated with AA, ACh, interferon pathways, and Covid-19 biomarkers (Supplementary Appendix III).

**Figure 5.**
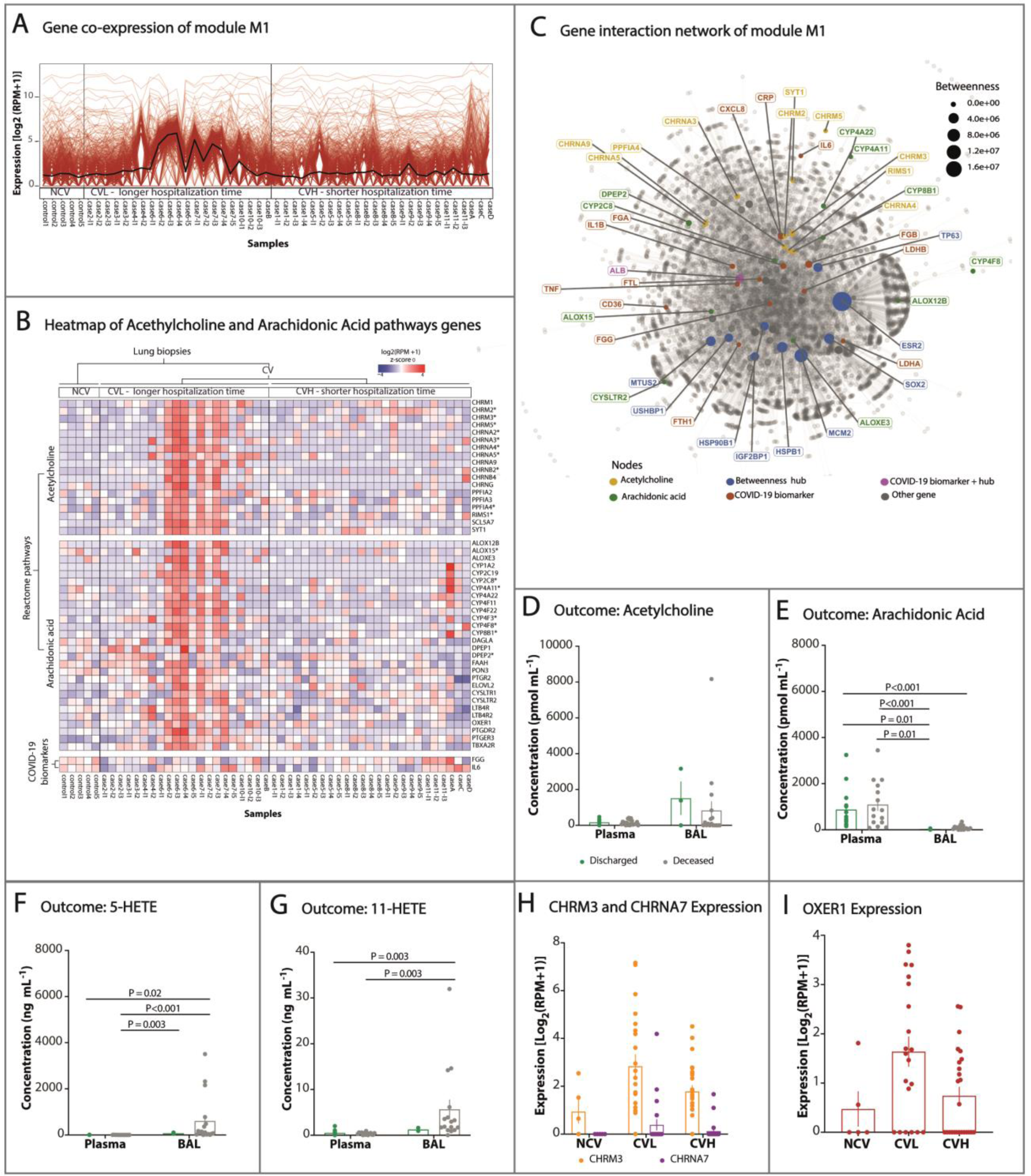
Re-analysis of transcriptome data reinforced the correlation between acetylcholine and arachidonic acid to Covid-19 severity and mortality. (A) Gene coexpression profile from CVL, CVH, and NCV samples in principal module M1 (1047 genes, red lines) and mean gene expression (black line) (n=51). Genes identified as being associated with AA and ACh were only found in module M1; ten are related to cholinergic receptors and the ACh release cycle and eight to AA metabolism along with several biomarkers related to Covid-19 severity (Supplementary Appendix II; Table S4). (B) Heatmap of normalized gene expression related to the ACh and AA pathways and Covid-19 biomarkers (n=51). Almost all coexpressed genes of the ACh and AA pathways were upregulated in the CVL versus CVH group, including twelve cholinergic receptors, six ACh release cycle genes, eighteen AA production and metabolism pathway genes, and nine eicosanoid receptors (Figure S6A; Supplementary Appendix III). Approximately 36% of these genes were also upregulated in the CVL versus NCV group but not in the CV versus NCV group (n=51); among them, elongation of very long-chain fatty acid protein 2 (ELOVL2) is involved in linoleic acid metabolism (Figure S6A; Supplementary Appendix III). Cases 3, 9, and 11 were glucocorticoid-treated patients. (C) First-order gene interaction network of the M1 module containing differentially expressed genes related to ACh (brown), AA (green), Covid-19 biomarkers (red), betweenness hubs (blue), and albumin genes (magenta – Covid-19 biomarker and hub). Ten DEGs related to ACh and ten DEGs of AA populated the biological network and are connected to thirteen genes involved in the pathophysiology of Covid-19 and CD36. ACh- and AA-related genes showed relatively low values of centrality metrics and may be under the action of some hub genes, such as oestrogen receptor II (ESR2), insulin-like growth factor II mRNA binding protein (IGF2BP1), albumin (ALB), and others (Supplementary Appendix IV). (D) Plasma and BAL levels of ACh (n=52). ACh levels in BAL fluid exhibited a tendency towards increase compared to plasma samples from deceased or discharged patients. (E) AA (n = 48) and AA concentrations were higher in plasma than in BAL in both patient groups. (F) 5-HETE (n = 48) and (G) 11-HETE (n=47) in several/critical patients according to outcome (discharged or deceased). 5-HETE and 11-HETE levels were only significantly higher in the BAL of deceased patients. (H) Transcription levels of cholinergic muscarinic M3 receptor (CHRM3) and cholinergic receptor nicotinic alpha 7 (CHRNA7) (n=51) and (I) oxoeicosanoid receptor 1 (OXER1) (n=51). A unique gene expression profile in lung samples from Covid-19 patients who had died was characterized by high levels of pro-inflammatory cholinergic receptors (CHRM3, CHRNA3, and CHRNA5) ^9, 110^, low levels of anti-inflammatory cholinergic receptor (CHRNA7) ^113^, and high expression of oxoeicosanoid receptor 1 (OXER1) ^50^ which mediates the pro-inflammatory effects of eicosanoids (Figures S6B, S6C). Other eicosanoid and cholinergic receptors were also differentially expressed in CVL patients compared to the CVH or NCV group (Figures S6A, S6B, S6C). Finally, transcriptome analysis of fifteen Covid-19 and five non-Covid-19 samples indicated a correlation between ACh and AA genes and Covid-19 severity in some CVL patients. Significant differences in BAL/plasma levels of mediators were set at P< 0.05 according to the Kruskal-Wallis test followed by Dunn’s posttest. *present in module M1. Abbreviations: Covid-19 low viral load (CVL) - Covid-19 high viral load (CVH) - non-Covid-19 (NCV).

## Discussion

A consensus is building around the fatal effects of SARS-CoV-2 infection, which is increasingly believed to cause death as a result of systemic hyperinflammation and multi-organ collapse^32^, secondary to systemic cytokine storms ^33–35^. However, few studies have compared pulmonary and systemic inflammation ^36^ or considered the contribution of eicosanoids and neurotransmitters to these effects. In our study, we hypothesised that, in Covid-19, pulmonary cells and leukocytes, in addition to cytokines, release eicosanoids and ACh, mediating local and systemic manifestations. In patients with severe/critical SARS-CoV-2 infection, we found that ACh, 5-HETE, 11-HETE, and cytokines were more abundant in the lung than under systemic conditions. In contrast, only the levels of AA were found to be higher in the circulation than in the BAL fluid. Interestingly, in patients with severe/critical disease who were treated with corticosteroids, only ACh was inhibited.

In our study, we compared the lung and systemic responses in association with the lung transcriptome and demonstrated a robust correlation between lipid mediators, neurotransmitters, and their receptors in SARS-CoV-2 infection. However, contrary to what has been suggested by previous studies ^37, 38^, only small amounts of eicosanoids were found in the plasma of patients with severe/critical disease, with significant differences observed only in AA, 5-HETE, and 11-HETE. Interestingly, the levels of 5-HETE and 11-HETE were found to be remarkably higher in BAL, as well as AA in plasma, suggesting that AA and its metabolites mediate responses to Covid-19. 5-HETE induces neutrophil recruitment ^39^, pulmonary oedema ^40^, and ACh release ^30^. It is released by human neutrophils, and its esterified form promotes IL-8 secretion ^31^. Unlike the esterified form ^31^, free-5-HETE did not inhibit NETs formation, a key event in Covid-19 ^41^. Remarkably, 11-HETE originating from monocytes/macrophages ^42^, endothelial cells ^43^, and platelets ^44^ is induced by hypoxia ^45^, IL-1β ^46^, contributes to ACh functions ^43^, and inhibits insulin release ^47^. The involvement of AA was confirmed by bioinformatic analyses that identified the expression or upregulation of genes related to AA metabolism and eicosanoid receptors in some lung biopsies. These included the OXER1 gene, which encodes a receptor for AA and 5-HETE ^48^ which mediates neutrophil activation/recruitment ^49, 50^. Our metabolomic analysis also showed an increase in the plasma AA and linoleic acid levels, similar to a plasma lipidome performed by Schwarz et al., suggesting a strong correlation between lipid mediators and Covid-19 severity ^51^. Accordingly, the ELOVL2 gene, which is involved in linoleic acid metabolism and AA synthesis ^52^, was upregulated in lung biopsies.

The RNA expression of ALOX5, an enzyme that participates in lipid mediator production, is upregulated in some immune cell types from severe Covid-19 patients ^53^, and has been detected in the lung biopsies of deceased Covid-19 patients ^18^. In contrast to our lung transcriptome re-analysis findings, the expression of cytochrome p450 (CYP) enzymes, which are also involved in lipid mediator generation, was not detected in the peripheral blood mononuclear cells (PBMC) transcriptome re-analysis from severe Covid-19 patients ^53^. One possible explanation for this contradiction is that Covid-19 is a heterogeneous illness composed of distinct tissue gene expression associated with Covid-19 severity and different immunopathological profiles in infected tissues ^18, 54^. This immunological heterogeneity could be related to two types of evolution of severe Covid-19 patients, one associated with a high viral load and susceptibility to SARS-CoV-2 infection, a shorter hospitalisation time, and exudative diffuse alveolar damage, and another associated with a low or undetectable viral load, a mixed lung histopathological profile, and a longer hospitalisation time associated with non-homeostatic pulmonary inflammation ^18, 54, 55^. In this context, we found that the expression levels of some lipid mediators and ACh and AA pathway genes varied in the plasma and BAL samples of severe/critical patients, which were found to be preferentially activated in the lungs of deceased Covid-19 patients with low viral loads, long hospitalisation times, and damaging lung inflammation.

Nevertheless, it remains unclear whether AA and linoleic acid contribute to host protection ^56^ and tissue damage, or whether they represent a viral escape mechanism. AA is known to directly interact with the virus, reducing its viability and inducing membrane disturbances, disfavouring SARS-CoV-2 entry ^57–59^. On the other hand, reduced plasma AA levels may be associated with lung injury and poor outcomes in Covid-19 infection ^60^. Within this context, Shen and et al. found lower concentrations of AA in survivors of severe Covid-19 infection ^37^. However, in the present study, the levels of AA were found to be increased in the plasma of patients who died from severe Covid-19, suggesting that the potential benefits of AA are overcome by a higher production of its proinflammatory metabolites, 5-HETE and 11-HETE. Interestingly, in our cohort, AA, 5-HETE, and 11-HETE were not altered in the plasma or BAL fluid of patients with severe/critical Covid-19 infection who had received glucocorticoid treatment. This may be explained by the fact that AA, in addition to calcium-activated-PLA2 ^61^ also originates from glucocorticoid-insensitive phospholipase ^62^, adipocyte destruction ^63^, linoleic acid ^64^, or the degradation of endocannabinoids by FAAH ^65^, a macrophage enzyme detected in SARS-CoV-2-infected patients ^66^. Notably, some CVL Covid-19 patients, regardless of whether they were treated or not with glucocorticoids, showed transcript expression of MGLL and NAAA, as well as an up-regulation of FAAH and all of the enzymes involved in the production of AA from endocannabinoid ^67^. In addition, we detected other DEGs in biopsy samples (glucocorticoid-treated versus non-treated) associated with AA, ACh, interferon pathways, and Covid-19 biomarkers. Glucocorticoids have been used widely to reduce the morbidity and mortality rates of Covid-19 patients, but have not been effective for all patients ^68^. The mortality rate for glucocorticoid-treated hospitalised Covid-19 patients from our cohort was higher than that reported in the RECOVERY trial and similar to that described in the CoDEX trial conducted in Brazil ^69^. High mortality rates could be associated with several factors, including a low mean PaO2:FiO2 ratio and overloaded public health systems in countries with limited resources, such as Brazil ^69^ and as well as glucocorticoid dose, initiation, and duration of therapy ^70, 71^. In addition, the precise threshold at which a patient should be treated with glucocorticoids, that is to avoid the manifestation of adverse effects associated with comorbidities and inefficient clearance of SARS-CoV-2, remains unclear ^72^. However, a delayed start of glucocorticoid therapy could result in a lack of response due to patients reaching a point of no return, as suggested previously by our research group with regards to scorpion poisoning, which triggers a sterile inflammatory process ^3^. On the other hand, considering that Covid-19 is a non-sterile hyperinflammatory disease, the administration of glucocorticoids is more effective after the initial phase in which hospitalised patients have low or undetectable viral loads ^18, 68^. Our data suggest that SARS-CoV-2 infection activates the glucocorticoid-insensitive release of AA metabolites associated with Covid-19 severity. Hence, lipid mediator production pathways could be important molecular targets for Covid-19 treatment, as suggested by other researchers ^53^. Accordingly, we suggested that, in order to improve the benefits of glucocorticoid therapy in hospitalized patients, who show lower or undetectable viral loads, patients should be treated as soon as possible in combination with AA metabolism inhibitors.

CD36 is expressed in several cells ^73, 74^, where it induces Ca^++^ mobilisation, cell signalling, LTB4 production, and cellular fatty acid uptake ^75–78^. The maintenance of high free-AA levels in patients with severe/critical glucocorticoid-treated disease may result from the reduction of CD36 on macrophage membranes. Intriguingly, CD36 is a gustatory lipid sensor ^79^, whose deficit in cell membranes may account for the symptoms of ageusia, anosmia, diarrhoea, hyperglycaemia, platelet aggregation, and cardiovascular disturbances experienced by Covid-19 patients ^75–77^. CD36 is a substrate of matrix metalloproteinase-9 (MMP-9) and disintegrin metalloproteinase domain-containing protein 17 (ADAM17) ^80, 81^. Decreased CD36 expression in Covid-19 patients most likely results from the action of these enzymes. MMP-9 is produced by neutrophils ^82^, induced by TNF-α ^83^, unaffected by glucocorticoids, and associated with respiratory syndrome in Covid-19 ^84^. ADAM17 has been described as a target for Covid-19 treatment ^85^.

In agreement with previous reports ^1, 34, 86, 87^, in the present study, high concentrations of IL-6, IL-8, and IL-10 were detected, along with insignificant levels of IL-1β in plasma. In contrast, the inflammatory cytokine TNF-α was found at the lowest concentration in BAL fluid, and insignificant levels were detected in the blood. The low levels of TNF-α production are correlated with the decreased counts of intermediate monocytes in the blood and BAL, which are major sources of this cytokine ^88^. As expected, in the BAL from patients with severe/critical Covid-19, the levels of IL-8, IL-6, IL-1β, TNF-α, and IL-10 were significantly higher compared with the plasma of all participants, or when paired with patients’ own plasma. Notably, in BAL, the most abundant cytokine was IL-8, which is produced by neutrophils ^89^, lung macrophages ^90^, and bronchial epithelial cells ^10^, and is induced by 5-HETE ^31^ and ACh ^10^. Interestingly, BAL fluid from non-Covid-19 patients also presented a high concentration of cytokines that did not differ from that of patients with Covid-19. This suggests that cytokine storms are not a key differential factor in this disease. Contrary to our expectations, glucocorticoids did not alter cytokine production (Table S7). As a result, we suggest that, regardless of whether Covid-19 is treated with glucocorticoids, cross-talk occurs between cytokines, lipid mediators, and ACh, as previously reported ^2, 3, 91^. IL-1β induces 11-HETE ^46^; arterial relaxation induced by ACh is mediated by 11-HETE and is inhibited by indomethacin ^43^. In our studies on scorpion envenomation, ACh release was found to be mediated by PGE2-induced by IL-1β and inhibited by indomethacin ^3^. In the present study, comparing AA, 5-HETE, 11-HETE, cytokine, and receptor expression demonstrated the absence of glucocorticoid effects. These results, in addition to the results of other studies from our laboratory ^3^, suggest that glucocorticoids did not have an effect in our cohort. This was most likely due to the fact that treatment was started late, namely after inflammation has been triggered by infection. In this context, Covid-19 patients could had already reached the point of no return, similar to what has been previously described in scorpion envenomation ^3^.

As predicted by Virgilis and Giovani ^92^, neurotransmitters are produced in Covid-19. ACh induces mucus secretion, bronchoconstriction, lung inflammation and remodelling ^9^, cardiac dysfunction in scorpionism ^3^, NETs formation ^93^, IL-8 release ^10^, thrombosis ^94^, and obesity-related severity ^95^. In addition to the nervous system ^4^, ACh is also produced by pulmonary vessels ^96^, airway epithelial cells ^6^, and immune cells ^5^. When binding to anti-inflammatory neural nicotinic receptors induces AA release ^97^, it inhibits nicotine receptors ^97^, which are highly expressed in lungs ^98^. In our cohort, a correlation between the AA and ACh levels and the severity of Covid-19 infection was observed, which was reinforced by the results of our bioinformatics study. Our lung transcriptome re-analyses showed that ACh release cycle genes were activated, including acetylcholine-synaptic release (synaptotagmin 1) and neuronal choline transporter (Solute Carrier Family 5 Member 7, SLC5A7). Accordingly, SLC5A7 gene expression was higher in the lung of patients who died from Covid-19 infection than patients who survived ^99^. Because this gene mediates the translocation of choline into lung epithelial cells ^100^ and macrophages ^101^, the production of ACh in BAL may also depend on non-neuronal cells. Indeed, the ACh repressor gene in non-neural cells (RE1 Silencing Transcription Factor) is reduced in the lungs of patients who died of Covid-19 ^99^. The activation of T-lymphocyte EP4 induces the release of ACh ^102^, suggesting that AA metabolites contribute to the release of ACh from lung and immune cells. As expected, ACh production in patients with severe/critical disease was found to be inhibited by glucocorticoids, since this drug blocks ACh production by lung epithelial cells ^103–105^. Patients with Covid-19 (mostly methylprednisolone-treated patients) showed lower plasma levels of choline and higher plasma levels of phosphocholine ^37^. In addition, a distinct pattern of ACh receptor mRNA expression was found in patients who died of Covid-19 (mainly in the CVL-lengthy hospital stay group). This profile is characterised by low levels of expression of the nicotinic receptor encoded by cholinergic receptor nicotinic alpha 7 subunit gene (CHRNA7) and increased levels of expression of two nicotinic and one muscarinic receptor in deceased-CVL compared to deceased non-Covid-19 patients. These proteins are encoded by the cholinergic nicotinic alpha 3 subunit gene (CHRNA3), the cholinergic nicotinic receptor 5 subunit gene (CHRNA5), and the muscarinic cholinergic receptor 3 gene (CHRM3), respectively. These three upregulated genes were also co-expressed and associated with pulmonary inflammatory disorders ^9, 106–110^. Notably, SARS-CoV-2 spike glycoprotein interacts with the α7 nicotinic acetylcholine receptor, which may compromise the cholinergic anti-inflammatory pathway^111^.

The ACh-nicotinic receptor, encoded by CHRNA7, is expressed in various lung cells and macrophages ^112^. It displays extensive anti-inflammatory activities, including the inhibition of pro-inflammatory cytokine production ^113^, the recruitment of neutrophils ^114^, and the reduction of CD14 expression in human monocytes ^115^. The increased degrees of inflammation in the BAL fluid of patients with severe/critical Covid-19 may be associated with the lower levels of anti-inflammatory CHRNA7 expression. However, a reduction in monocyte CD14 may result from high levels of IL-6 production ^116^, since the levels of CHRNA7 expression were very low. Interestingly, the ACh-M3 receptor in immune cells has been found to be up-regulated by ACh, which mediates its pro-inflammatory actions, including the production of IL-8 and the recruitment of neutrophils ^117^. Furthermore, the interaction between ACh and its receptor triggers the AA-derived release of eicosanoids, including 5-HETE ^97, 118, 119^. Remarkably, vitamin D modulates the ACh-M3 receptor ^120^, which may explain its beneficial effects in the prevention of Covid-19 ^121^. Based on the anti-inflammatory effects of nicotinic receptors and the downregulation of the SARS-CoV-2 receptor ACE2 promoted by nicotine, therapies involving nicotinic receptors have been proposed to treat Covid-19 infection ^122, 123^. In fact, based on these findings, an acetylcholinesterase inhibitor ^124, 125^ and a nicotinic receptor agonist have been tested ^126, 127^. However, recent studies have demonstrated that nicotine and smoking increase ACE2 receptor density ^128, 129^, and that the potential beneficial effects of nicotine were restricted to a small group of individuals ^130^. Our findings provide a warning that precautions should be taken when considering the therapeutic use of nicotinic agonists, since patients with severe/critical Covid-19 were found to release high amounts of ACh, in addition to showing increased levels of M3-receptor expression. Due to the antiviral and anti-inflammatory effects of AA, it has been proposed to treat Covid-19 patients ^60^. Nevertheless, AA may be not be an adequate therapeutic target, based on both our findings and previous studies ^131^, which have shown that AA can favour hyperinflammation and lethality in Covid-19 infection. In conclusion, ACh, AA, 5-HETE, and 11-HETE mediate the innate immune response to SARS-CoV-2 and may define the outcome of infection.

Covid-19 severity can be associated with disruption of homeostatic lipidome and metabolic alterations and this phenomenon may be influenced by comorbidities/risk factors related to the infection. Our findings demonstrated that (i) high plasma levels of ACh and lipid mediators positively correlated with Covid-19 severity; and (ii) only hypertension and/or age were confounding variables for analyzing the association of high plasma levels of AA, 5-HETE, and ACh with disease severity (Table S7). Similarly, the correlation between altered plasma profile of lipid mediators and Covid-19 severity is associated with selective comorbidities – mainly high body mass index (BMI) – but poorly associated with gender, advanced age, and diabetes. However, the correlation between altered AA and/or 5-HETE levels and disease severity is associated with male gender, hypertension, and heart disease, but not BMI^53^.

We also examined whether glucocorticoid-therapy interfered with the potential effect of some confounding variables on the correlation between high ACh levels and Covid-19 severity in severe/critical patients. Despite the relatively underrepresented samples from non-glucocorticoid treated patients, no confounding variable significantly affected the correlation between plasma and BAL ACh levels and Covid-19 severity in severe/critical patients treated or not with glucocorticoids (Table S8). The altered plasma and BAL levels of the lipid mediators AA, 5-HETE, and 11-HETE in severe/critical patients were associated with the disease severity but not with the confounding variables tested (Table S9). Altogether, the findings here reported suggest that age and/or hypertension are significant confounding variables for analyzing the association between increased levels of cholinergic and lipid mediators and Covid-19 severity. Also, the confounding variables tested probably did not modify the inhibitory action of glucocorticoids on plasma and BAL ACh levels in severe/critical patients.

To the best of our knowledge, this study is the first to demonstrate that the lung inflammatory process and poor outcomes of patients with Covid-19 infection are associated with high levels of lipid mediators and ACh, produced via a partially glucocorticoid-insensitive pathway. Glucocorticoid therapy was found to lower only the levels of ACh. Thus, to improve the benefits of glucocorticoid therapy, we suggest that treatment in hospitalised patients be started early and be preferentially administered to patients with low or undetectable viral loads and harmful lung inflammation in combination with AA metabolism inhibitors.

## Conclusions

We demonstrate for the first time that the lung inflammatory process and worse outcomes in Covid-19 are associated with lipid mediators and ACh, which are produced through a partially glucocorticoid-insensitive pathway. To improve the benefits of glucocorticoid therapy, we suggest that it should be started early in severe/critical hospitalized patients and in combination with AA metabolism inhibitors.

## Supporting information

Supplementary Appendix III

ETHICAL APPROVAL STATEMENT

## Data Availability

All data can be requested by

## Acknowledgements

The authors thankfully acknowledge Innovation and Technology Park (Supera), the healthy-participants joining as controls and the positive COVID-19 patients as well as their families. We grieve for all patients who lost their lives as a result of this pandemic, including those who provided us with samples to be able to answer scientific questions and contribute to humanity’s eradication of this disease. We also thank Fabiana Rossetto de Moraes, B.Sc., for the cytometry analysis, Caroline Fontanari, M.Sc., for laboratory and technical support, the ICU team, and all hospital professionals, especially the technicians, nurses, physiotherapists, and biomedical personnel, who collaborated on this work through Hospital Santa Casa de Misericórdia in Ribeirão Preto and Hospital São Paulo in Ribeirão Preto. We are grateful for the indispensable contribution of the Ribeirão Preto Municipal Health Department and the employees of the Serviço de Análises Clínicas (SAC) of the Faculdade de Ciências Farmacêuticas de Ribeirão Preto, USP. We also thank Professors Victor Hugo Aquino Quintana, Ph.D., Márcia Regina von Zeska Kress, Ph.D., and Marcia Eliana da Silva Ferreira, Ph.D. for sharing the BSL2 viral laboratory.

## Additional Information

Extended data available in Supplementary Appendixes:

- Supplementary Appendix I - Pathways and list of corresponding genes related to acetylcholine and arachidonic acid observed in Reactome pathways and Covid-19 biomarkers.
- Supplementary Appendix II - Report of CEMITool results for the gene co-expression modular analysis of lung samples from biopsies of Covid-19 and non-Covid-19 patients.
- Supplementary Appendix III - Differential gene expression analysis results between Covid-19-death groups (CV, NCV, CVL and CVH) and Covid-19 samples from patients who underwent glucocorticoid treatment (GC) and not (non-GC).
- Supplementary Appendix IV - Betweenness and degree values of genes from biological network constructed based on the principal co-expression module M1.

## Funding

Fundação de Amparo a Pesquisa do Estado de São Paulo (FAPESP): #2020/05207-6, #2014/07125-6 and #2015/00658-1 for **L.H.F**.; #2020/08534-8 for **M.M.P**.; #2018/22667-0 for **C.O.S**.**S.;** #2020/05270-0 for **V.D.B**. Additional support was provided by the National Council for Scientific and Technological Development (CNPq), the Coordination for the Improvement of Higher Educational Personnel (CAPES-Finance Code 001)), and Pró-Reitora de Pesquisa da Universidade de São Paulo, grant-USP-VIDA.

## Conflict of Interest Statement

The authors declare that this research was performed without conflicts of interest or commercial or financial gains.

## Supplementary Tables

**Table S1.**
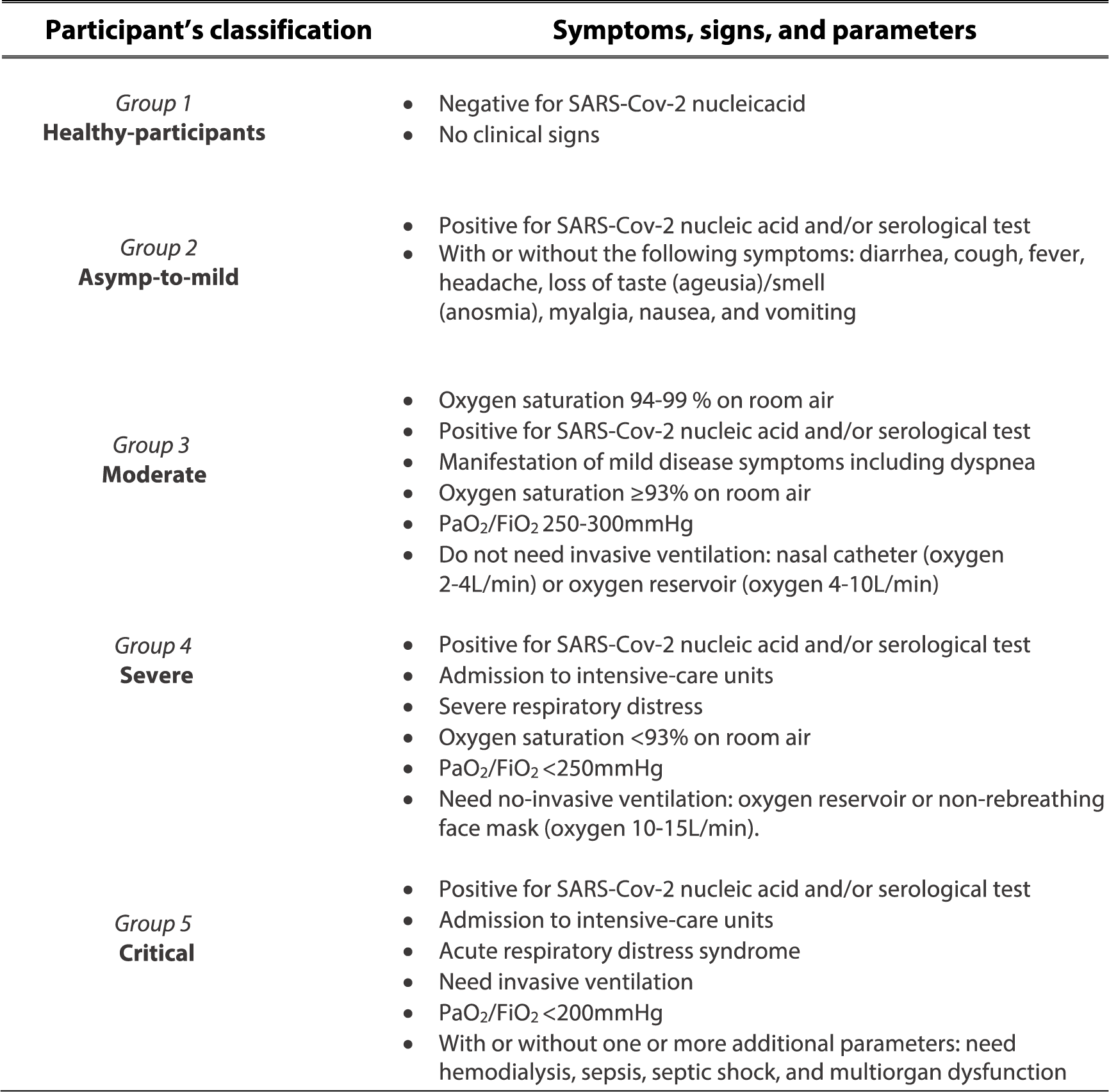
Classification of study participants. The participants were classified into five clinical groups (G1–G5): healthy participants (G1) and Covid-19 patients (G2 to G5), based on the severity of disease, clinical parameters, patient’s management, and laboratory findings, following the recommendations from WHO ^132–137^. These classifications were used to define the scale of the clinical progression of patients. The inclusion criteria were as follows: (i) signed informed consent form; (ii) fit into one of the five clinical groups; (iii) healthy participants must be negative for SARS-CoV-2 nucleic acid; (iv) asymptomatic or symptomatic participants must be positive for SARS-CoV-2 nucleic acid and/or anti-SARS-CoV-2 antibody; (v) age ≥12 years. Participants from groups G1, G2, and G5 were 16 years old or older, while the participants from groups G3 and G4 were 12 years old or older. Pregnancy was the only exclusion criterion for healthy participants (G1). Abbreviations: FiO2, fraction of inspired oxygen; PaO2, partial pressure of oxygen.

| Participant's classification | Symptoms, signs, and parameters |
| --- | --- |
| *Group 1*<br>**Healthy-participants** | Negative for SARS-Cov-2 nucleic acid<br>No clinical signs |
| *Group 2*<br>**Asymp-to-mild** | Positive for SARS-Cov-2 nucleic acid and/or serological test<br>With or without the following symptoms: diarrhea, cough, fever, headache, loss of taste (ageusia)/smell (anosmia), myalgia, nausea, and vomiting |
| *Group 3*<br>**Moderate** | Oxygen saturation 94-99 % on room air<br>Positive for SARS-Cov-2 nucleic acid and/or serological test<br>Manifestation of mild disease symptoms including dyspnea<br>Oxygen saturation $\geq 93\%$ on room air<br>$PaO_2/FiO_2$ 250-300mmHg<br>Do not need invasive ventilation: nasal catheter (oxygen 2-4L/min) or oxygen reservoir (oxygen 4-10L/min) |
| *Group 4*<br>**Severe** | Positive for SARS-Cov-2 nucleic acid and/or serological test<br>Admission to intensive-care units<br>Severe respiratory distress<br>Oxygen saturation $< 93\%$ on room air<br>$PaO_2/FiO_2 < 250$mmHg<br>Need no-invasive ventilation: oxygen reservoir or non-rebreathing face mask (oxygen 10-15L/min). |
| *Group 5*<br>**Critical** | Positive for SARS-Cov-2 nucleic acid and/or serological test<br>Admission to intensive-care units<br>Acute respiratory distress syndrome<br>Need invasive ventilation<br>$PaO_2/FiO_2 < 200$mmHg<br>With or without one or more additional parameters: need hemodialysis, sepsis, septic shock, and multiorgan dysfunction |

**Table S2.**
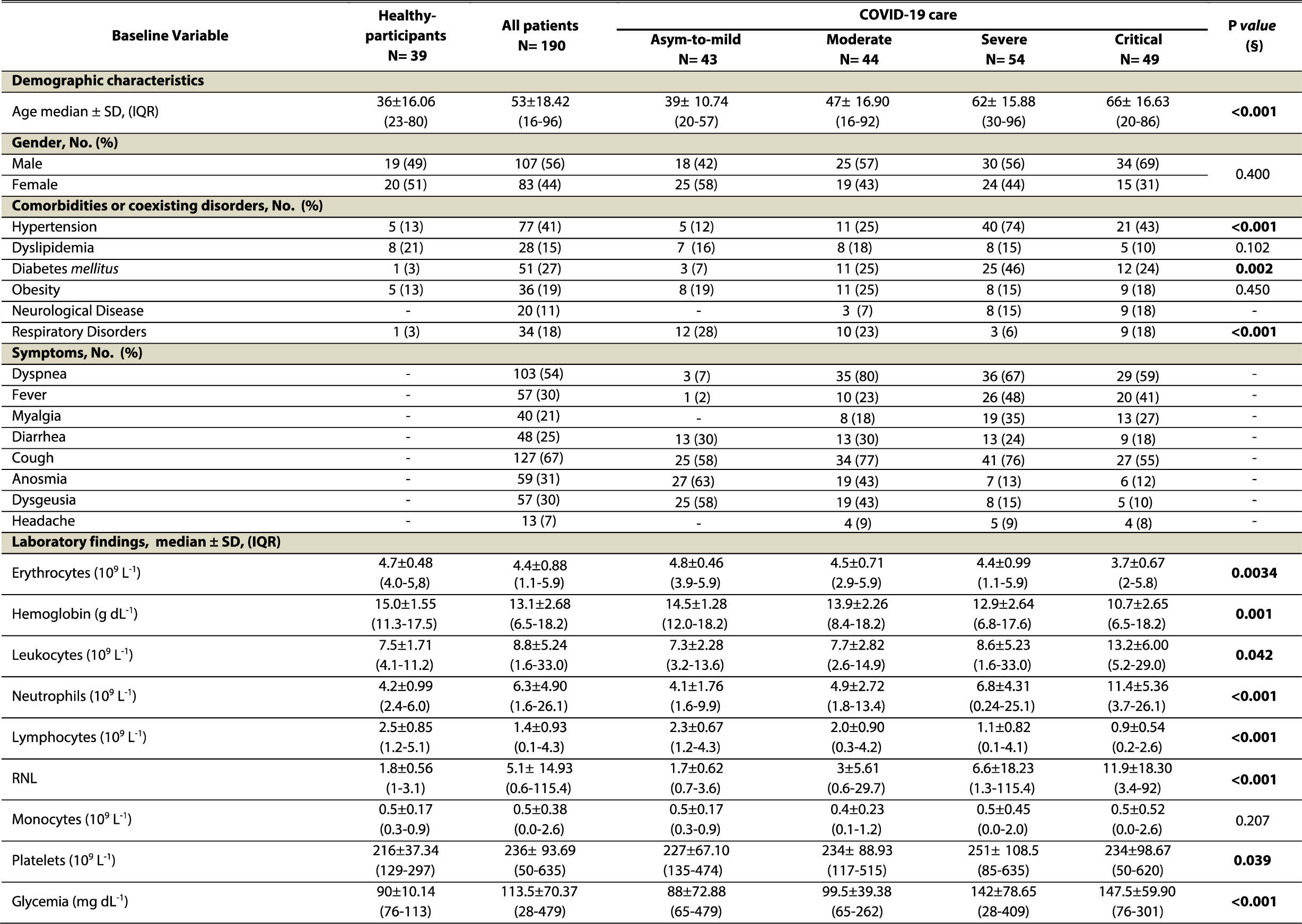
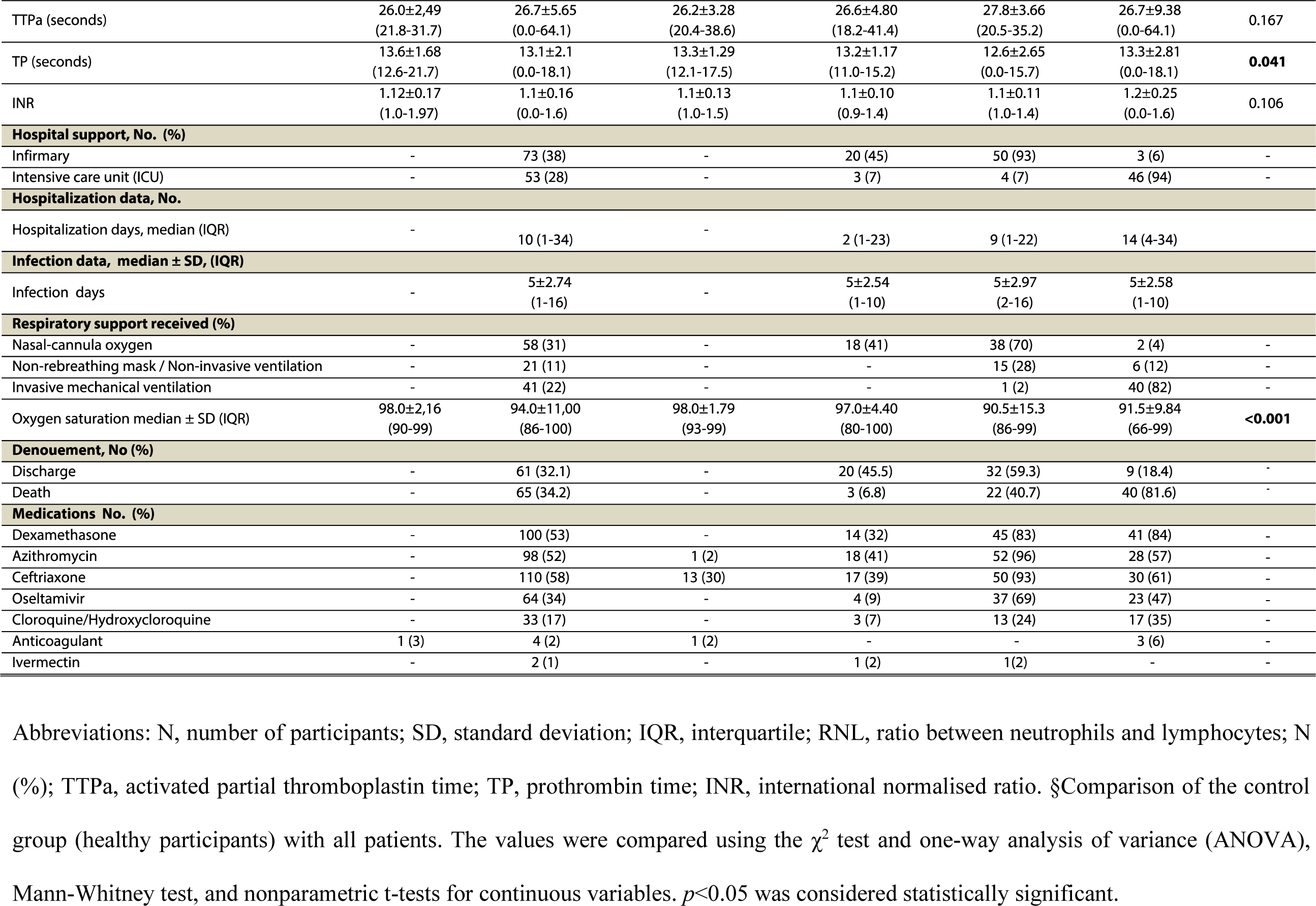
Data of demographic, clinical, and blood findings (n=229)

**Table S3.**
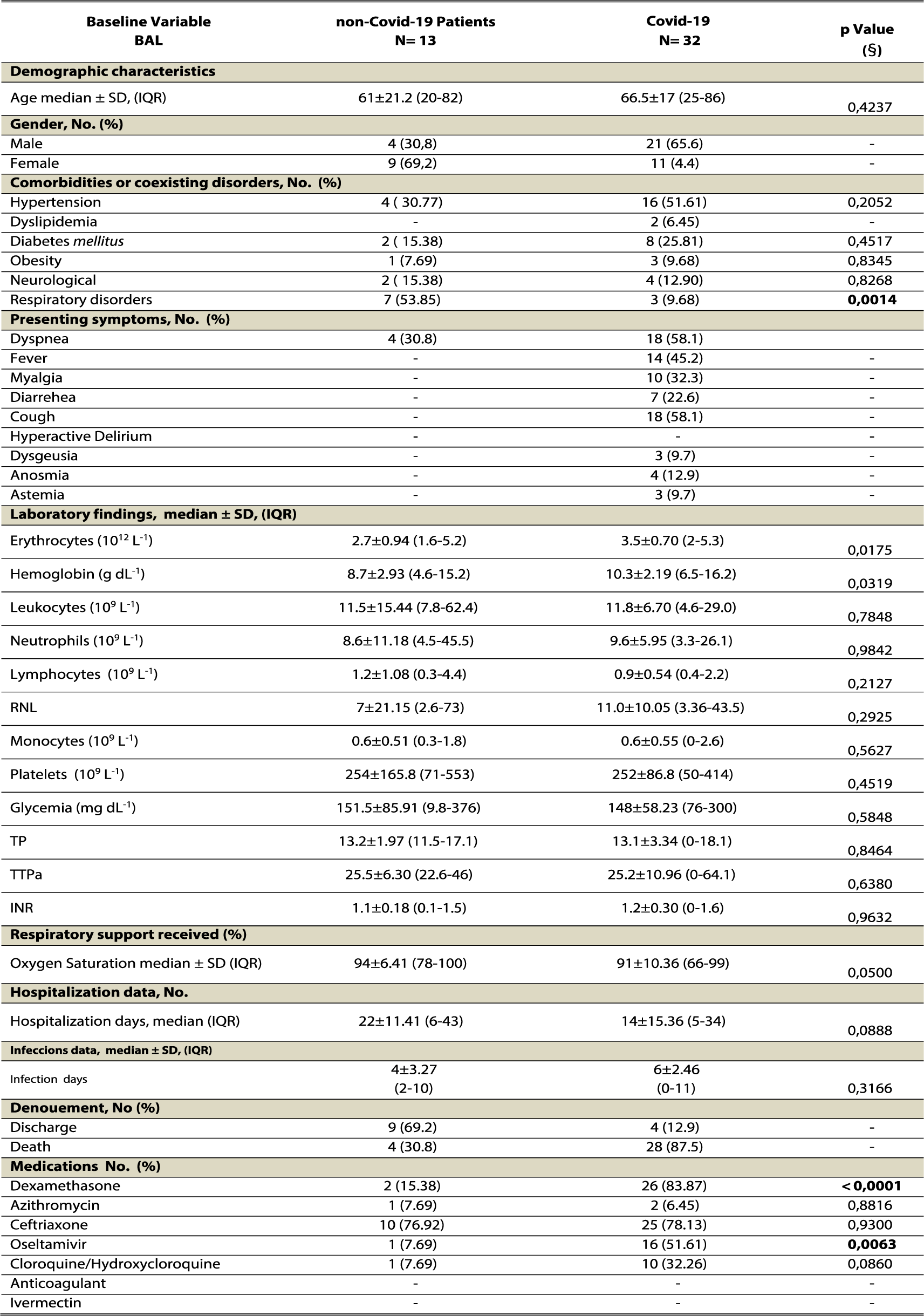
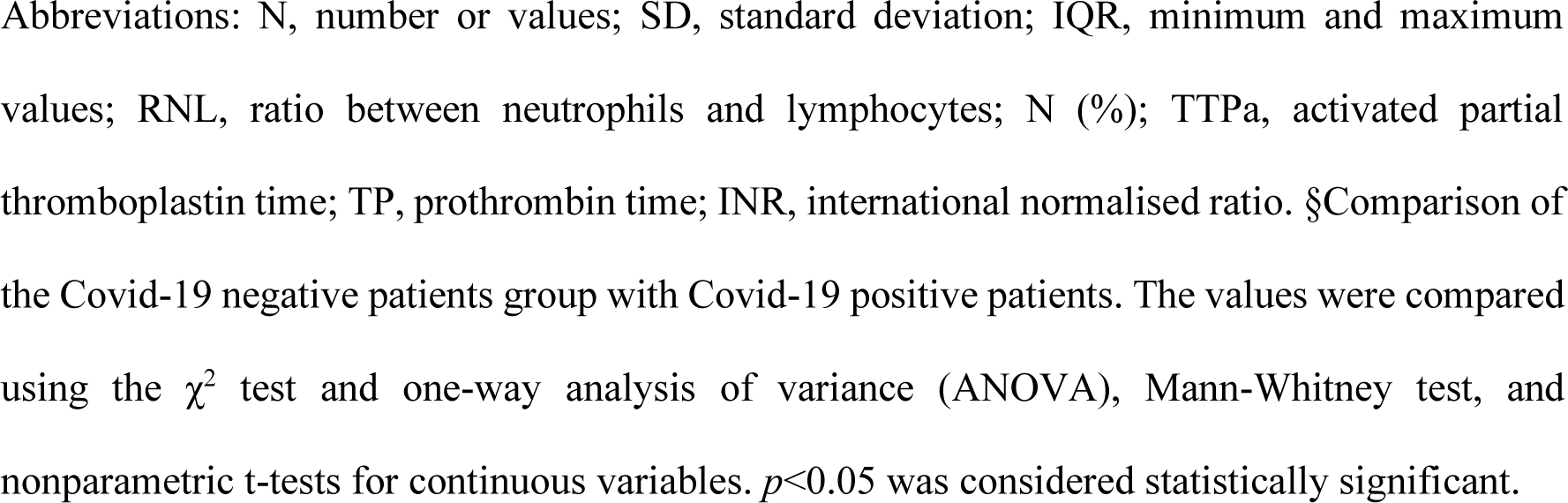
Demographic, clinical characteristics and blood findings data from severe/critical participants of which bronchoalveolar lavage (BAL) were collected (n=45)

| Baseline Variable<br>BAL | non-Covid-19 Patients<br>N= 13 | Covid-19<br>N= 32 | p Value<br>(§) |
| --- | --- | --- | --- |
| **Demographic characteristics** |  |  |  |
| Age median ± SD, (IQR) | 61±21.2 (20-82) | 66.5±17 (25-86) | 0,4237 |
| **Gender, No. (%)** |  |  |  |
| Male | 4 (30,8) | 21 (65,6) | - |
| Female | 9 (69,2) | 11 (4,4) | - |
| **Comorbidities or coexisting disorders, No. (%)** |  |  |  |
| Hypertension | 4 ( 30,77) | 16 (51,61) | 0,2052 |
| Dyslipidemia | - | 2 (6,45) | - |
| Diabetes *mellitus* | 2 ( 15,38) | 8 (25,81) | 0,4517 |
| Obesity | 1 (7,69) | 3 (9,68) | 0,8345 |
| Neurological | 2 ( 15,38) | 4 (12,90) | 0,8268 |
| Respiratory disorders | 7 (53,85) | 3 (9,68) | **0,0014** |
| **Presenting symptoms, No. (%)** |  |  |  |
| Dyspnea | 4 (30,8) | 18 (58,1) | - |
| Fever | - | 14 (45,2) | - |
| Myalgia | - | 10 (32,3) | - |
| Diarrhea | - | 7 (22,6) | - |
| Cough | - | 18 (58,1) | - |
| Hyperactive Delirium | - | - | - |
| Dysgeusia | - | 3 (9,7) | - |
| Anosmia | - | 4 (12,9) | - |
| Astemia | - | 3 (9,7) | - |
| **Laboratory findings, median ± SD, (IQR)** |  |  |  |
| Erythrocytes (10<sup>12</sup> L<sup>-1</sup>) | 2.7±0.94 (1,6-5,2) | 3.5±0.70 (2-5,3) | 0,0175 |
| Hemoglobin (g dL<sup>-1</sup>) | 8.7±2.93 (4,6-15,2) | 10.3±2.19 (6,5-16,2) | 0,0319 |
| Leukocytes (10<sup>9</sup> L<sup>-1</sup>) | 11.5±15.44 (7,8-62,4) | 11.8±6.70 (4,6-29,0) | 0,7848 |
| Neutrophils (10<sup>9</sup> L<sup>-1</sup>) | 8.6±11.18 (4,5-45,5) | 9.6±5.95 (3,3-26,1) | 0,9842 |
| Lymphocytes (10<sup>9</sup> L<sup>-1</sup>) | 1.2±1.08 (0,3-4,4) | 0.9±0.54 (0,4-2,2) | 0,2127 |
| RNL | 7±21.15 (2,6-73) | 11.0±10.05 (3,36-43,5) | 0,2925 |
| Monocytes (10<sup>9</sup> L<sup>-1</sup>) | 0.6±0.51 (0,3-1,8) | 0.6±0.55 (0-2,6) | 0,5627 |
| Platelets (10<sup>9</sup> L<sup>-1</sup>) | 254±165.8 (71-553) | 252±86.8 (50-414) | 0,4519 |
| Glycemia (mg dL<sup>-1</sup>) | 151.5±85.91 (9,8-376) | 148±58.23 (76-300) | 0,5848 |
| TP | 13.2±1.97 (11,5-17,1) | 13.1±3.34 (0-18,1) | 0,8464 |
| TTPa | 25.5±6.30 (22,6-46) | 25.2±10.96 (0-64,1) | 0,6380 |
| INR | 1.1±0.18 (0,1-1,5) | 1.2±0.30 (0-1,6) | 0,9632 |
| **Respiratory support received (%)** |  |  |  |
| Oxygen Saturation median ± SD (IQR) | 94±6,41 (78-100) | 91±10,36 (66-99) | 0,0500 |
| **Hospitalization data, No.** |  |  |  |
| Hospitalization days, median (IQR) | 22±11,41 (6-43) | 14±15,36 (5-34) | 0,0888 |
| **Infections data, median ± SD, (IQR)** |  |  |  |
| Infection days | 4±3,27<br>(2-10) | 6±2,46<br>(0-11) | 0,3166 |
| **Denouement, No (%)** |  |  |  |
| Discharge | 9 (69,2) | 4 (12,9) | - |
| Death | 4 (30,8) | 28 (87,5) | - |
| **Medications No. (%)** |  |  |  |
| Dexamethasone | 2 (15,38) | 26 (83,87) | **<0,0001** |
| Azithromycin | 1 (7,69) | 2 (6,45) | 0,8816 |
| Ceftriaxone | 10 (76,92) | 25 (78,13) | 0,9300 |
| Oseltamivir | 1 (7,69) | 16 (51,61) | **0,0063** |
| Cloroquine/Hydroxychloroquine | 1 (7,69) | 10 (32,26) | 0,0860 |
| Anticoagulant | - | - | - |
| Ivermectin | - | - | - |

Abbreviations: N, number or values; SD, standard deviation; IQR, minimum and maximum values; RNL, ratio between neutrophils and lymphocytes; N (%); TTPa, activated partial thromboplastin time; TP, prothrombin time; INR, international normalised ratio. §Comparison of the Covid-19 negative patients group with Covid-19 positive patients. The values were compared using the $\chi^2$ test and one-way analysis of variance (ANOVA), Mann-Whitney test, and nonparametric t-tests for continuous variables. $p < 0.05$ was considered statistically significant.

**Table S4.**
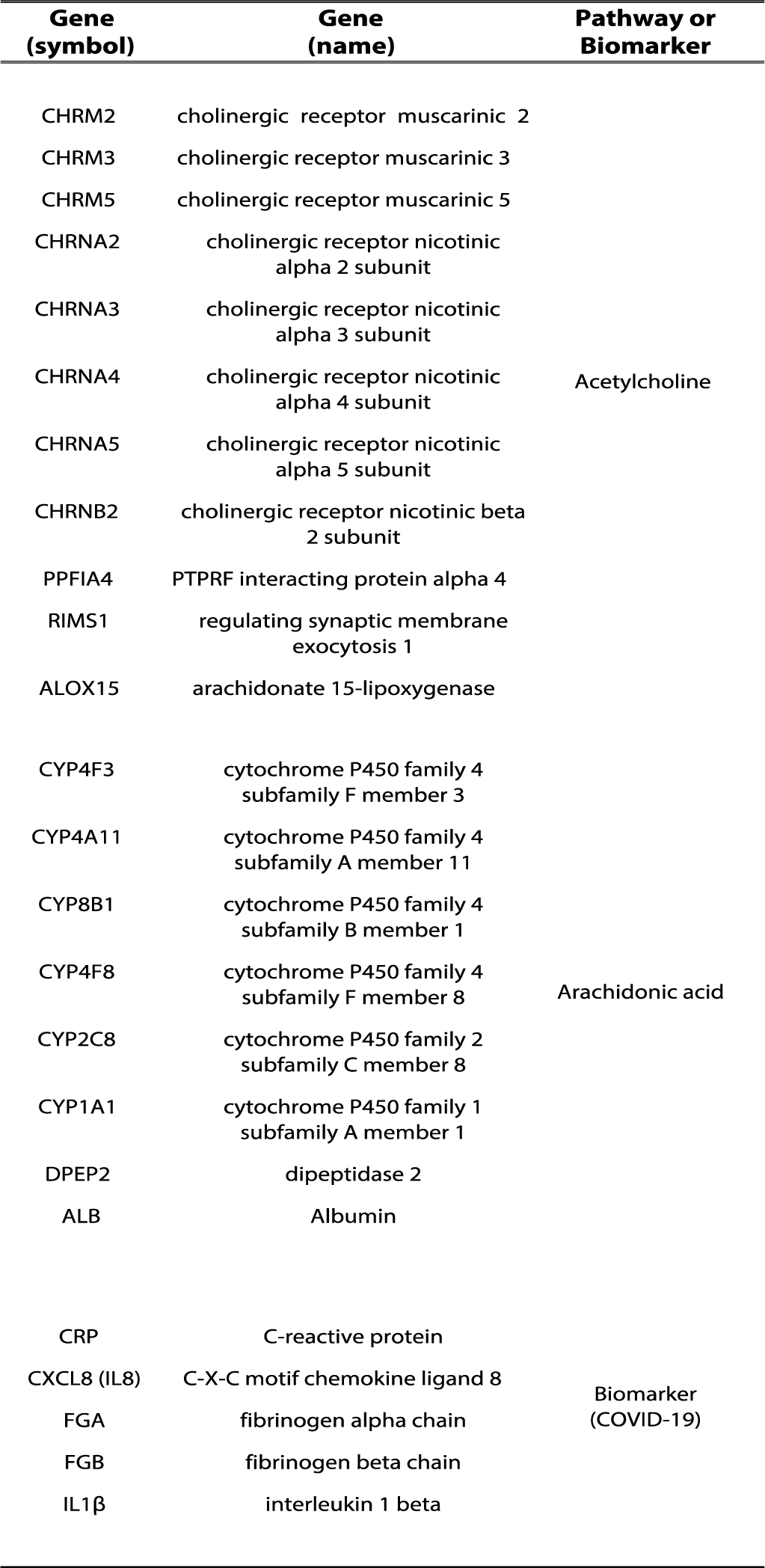
List of genes related to acetylcholine and arachidonic acid pathways and Covid-19 biomarkers founded in co-expression module M1. Co-expression analysis generated five different co-expression gene modules (M1 to M5) and only in M1 we identified genes that encoding proteins associated to cholinergic receptors (muscarinic and nicotinic), ACh release cycle, and AA metabolism. In addition, the M1 module also contains some biomarkers related to Covid-19 severity (albumin, C-reactive protein, IL-8, fibrinogen, and IL1β)^21,34,138,139^.

**Table S5.** Treatment of severe and critical Covid-19 patients with glucocorticoids does not alter the levels of plasmatic cytokines and circulating neutrophils and lymphocytes. Abbreviations: N, number of participants; SEM, standard error of the mean; IQR, interquartile; RNL, ratio between neutrophils and lymphocytes. §Comparison of the severe non-GC group with the severe GC group, and the critical non-GC group with the critical GC group. The values were compared using the χ^2^ test, one-way analysis of variance (ANOVA), Mann-Whitney test, and nonparametric t-tests for continuous variables. *p*<0.05 was considered statistically significant. As shown in the table, glucocorticoid therapy in severe Covid-19 patients did not show a statistically significant influence on the plasma levels of the cytokines evaluated in our study. The use of glucocorticoids and their impact on the production of inflammatory molecules is closely associated with several factors, such as the dose, duration, and time of initiation of therapy, since, in the critical stages of Covid-19, no beneficial effects of glucocorticoids have been observed in the inflammation control and patient outcome ^70, 71^.

|  | **Severe**<br>**non-GC**<br>**N=3** | **Severe**<br>**GC**<br>**N=39** | **P value**<br>**(#)** | **Critical**<br>**non-GC**<br>**N=4** | **Critical**<br>**GC**<br>**N=17** | **P value**<br>**(§)** |
| --- | --- | --- | --- | --- | --- | --- |
| **Parameters, median ± SEM, (IQR)** |  |  |  |  |  |  |
| **IL-8** | 31.7±12.6<br>(13.6-56.0) | 26.3±4.97<br>(2.8-186.4) | 0.454 | 26.86±6.5<br>(10.2-41.8) | 113.2±48.8<br>(13.1-661.9) | 0.144 |
| **IL-6** | 42.7±20.1<br>(4.80-73.2) | 54.9±14.19<br>(3.3-414.1) | 0.782 | 46.4±13.2<br>(24.7-81.7) | 317.7<br>±160.3<br>(11.3-2510) | 0.410 |
| **IL-1** | 2.5±2.5<br>(0.0-7.6) | 0.1±0.05<br>(0.0-1.7) | 0.104 | 0.9±0.9<br>(0.0-3.6) | 0.5±0.3<br>(0.0-3.8) | 0.874 |
| **TNF** | 0.0±0.0<br>(0.0-0.0) | 0.01±0.1<br>(0.0-0.3) | >0.999 | 0.8±0.8<br>(0.0-3.1) | 0.04±0.04<br>(0.0-0.7) | 0.191 |
| **IL-10** | 1.0±0.5<br>(0.4-2.1) | 2.7±0.4<br>(0.3-9.1) | 0.165 | 2.3±1.1<br>(0.8-4.3) | 7.5<br>±2.6<br>(0.1-45.1) | 0.229 |
| **Neutrophils** | 56.5±20.8<br>(15.0-77.5) | 80.5±1.7<br>(38.6-97.0) | 0.097 | 79.9±3.8<br>(73.1-89.0) | 85.6±1.5<br>(70.1-93.0) | 0.197 |
| **Lymphocytes** | 9.7±1.6<br>(8.0-13.0) | 12.0±0.9<br>(2.0-27.8) | 0.490 | 10.9±2.2<br>(5.0-14.7) | 7.9±1.0<br>(4.0-20.5) | 0.156 |
| **RNL** | 5.7±2.2<br>(1.9-9.4) | 10.2±1.6<br>(2.2-47.5) | 0.403 | 9.0±2.7<br>(5.1-16.6) | 13.3±1.5<br>(3.4-23.3) | 0.140 |

Abbreviations: N, number of participants; SEM, standard error of the mean; IQR, interquartile; RNL, ratio between neutrophils and lymphocytes. §Comparison of the severe non-GC group with the severe GC group, and the critical non-GC group with the critical GC group. The values were compared using the $\chi^2$ test, one-way analysis of variance (ANOVA), Mann-Whitney test, and nonparametric t-tests for continuous variables. $p < 0.05$ was considered statistically significant.

**Table S6.** Impact of glucocorticoid therapy in the outcome of hospitalized Covid-19 patients. Abbreviations: N, number of participants (%). Hospitalised Covid-19 patients from the moderate, severe, and critical groups were administered glucocorticoid (GC) therapy or not (non-GC) (methylprednisolone; range 40 to 500 mg/kg/day, or dexamethasone; range 1.5 to 6.0 mg/kg/day, by intravenous route). Most hospitalised patients were treated with glucocorticoids (moderate [69.6%], severe [87.0%], and critical [73.5%]). All moderate non-GC-patients were discharged, while GC patients had a mortality rate of 18.8%. In addition, the mortality rates for severe-GC (40.4%) and critical-GC (86.1%) patients were higher than those for moderate-CG patients. The discharge rates of non-GC patients vary according to the clinical categorization (100.0%, 57.1%, and 30.8% for moderate, severe, and critical, respectively). Hospitalised Covid-19 patients from our cohort showed fewer benefits of glucocorticoid therapy than those reported in the RECOVERY trial ^68^, but similar benefits to those described in the CoDEX trial conducted in Brazil ^69^. This reduction of glucocorticoid benefits could be associated with several factors, including a low mean PaO2:FiO2 ratio and an overload of the health systems of countries with limited resources, such as Brazil ^69^, as well as glucocorticoid dose, initiation, and duration of therapy ^70, 71^.

|  | Moderate<br>N=23 |  | Severe<br>N=54 |  | Critical<br>N=49 |  |
| --- | --- | --- | --- | --- | --- | --- |
| No, (%) | Discharge<br>N=20 | Death<br>N=3 | Discharge<br>N=32 | Death<br>N=22 | Discharge<br>N=9 | Death<br>N=40 |
| **non-GC** | 7 (35) | 0 (0) | 4 (12.5) | 3 (13.6) | 4 (44.4) | 9 (22.5) |
| **GC** | 13 (62) | 3 (100) | 28 (87.5) | 19 (86.4) | 5 (55.6) | 31 (77.5) |

Abbreviations: N, number of participants (%).

**Table S7.** Analysis of the potential interference of Covid-19-confounding variables on the correlation between high plasma levels of cholinergic and lipid mediators and Covid-19 severity. We analysed some confounding variables associated with Covid-19, such as comorbidities (Diabetes mellitus, hypertension, and obesity) and risk factors (advance age and gender) ^51, 140^, using data from participants whose plasma levels of lipid mediators (AA, 5-HETE, and 11-HETE) and ACh were measured. Lipid mediators data came from Covid-19 patients and heathy participants presented in Figure 1 (Panels: F, G, and H), while ACh data came from glucocorticoid-non-treated Covid-19 patients presented in Figure 4E. Age and hypertension or only age are potential confounding variables on the correlation between high plasma levels of lipid mediators or ACh and Covid-19 severity, because (i) age had significant Spearman’s correlation with AA (r=0.36; P=0.0042), 5-HETE (r=0.36; P=0.0042), and ACh (r=0.37; P = 0.0485); and (ii) hypertensive patients had significantly increased plasma levels of AA (*p*=0.0062 - Mann-Whitney test). §Kruskal-Wallis or Fisher’s tests were used to compare the differences between all clinical categories. Data are expressed as median (IQR - interquartile range) or number (% - percentages), and P < 0.05 was considered statistically significant.

| Confounding variables | Healthy participants | Asym-to-mild | Moderate | Severe | Critical | P value (\$) |
| --- | --- | --- | --- | --- | --- | --- |
| **Lipid mediators (AA, 5-HETE, and 11-HETE)** |  |  |  |  |  |  |
| No | 17 | 10 | 11 | 16 | 11 |  |
| Age, mean (IQR) | **33 (27-44)** | **33 (26-43)** | **39 (30-48)** | **57 (48-78)** | **71 (57-78)** | **<0.001** |
| Gender, No (%) Male |  |  |  |  |  | 0.795 |
| Female | 7 (41.2) | 5 (50.0) | 6 (54.5) | 7 (43.8) | 7 (63.6) |  |
| Diabetes, No (%) | 10 (58.8) | 5 (50.0) | 5 (45.5) | 9 (56.2) | 4 (36.4) |  |
| Hypertension, No (%) | 1 (5.9) | 1 (10.0) | 3 (27.3) | 6 (37.5) | 2 (18.2) | 0.183 |
| Obesity, No (%) | **2 (11.8)** | **0 (0.0)** | **0 (0.0)** | **12 (75.0)** | **6 (54.5)** | **<0.001** |
|  | 4 (23.5) | 2 (20.0) | 3 (27.3) | 2 (12.5) | 1 (9.1) | 0.7788 |
| **ACh** |  |  |  |  |  |  |
| No | 6 | 5 | 4 | 6 | 8 |  |
| Age, mean (IQR) | **30 (24-56)** | **42 (38-46)** | **48 (39-57)** | **75 (63-79)** | **65 (52-74)** | **0.026** |
| Gender, No (%) Male |  |  |  |  |  | 0.290 |
| Female | 4 (66.7) | 3 (60.0) | 2 (50.0) | 1 (16.7) | 6 (75.0) |  |
| Diabetes, No. (%) | 2 (33.3) | 2 (40.0) | 2 (50.0) | 5 (83.3) | 2 (25.0) |  |
| Hypertension, No. (%) | 0 (0.0) | 1 (20.0) | 1 (25.0) | 3 (50.0) | 1 (12.5) | 0.282 |
| Obesity, No. (%) | 2 (33.3) | 2 (40.0) | 1 (25.0) | 5 (83.3) | 3 (37.5) | 0.360 |
|  | 1 (16.7) | 1 (20.0) | 0 (0.0) | 0 (0.0) | 2 (25.0) | 0.767 |

**Table S8.** Analysis of the potential interference of Covid-19-confounding variables on the correlation between high ACh levels in severe/critical patients, treated or not with glucocorticoids, and Covid-19 severity. Here we used the same confounding variables associated with Covid-19 described on Table S7. This analysis was based on plasma and BAL cholinergic mediator (ACh) data from severe/critical Covid-19 patients, treated (GC) or not (non-GC) with glucocorticoids, and reported in Figure 4 (Panels: F and G). None of the confounding variables tested had significant potential to interfere with the glucocorticoid-therapy ability to reduce the high plasma and BAL ACh levels in severe/critical patients. §Mann-Whitney or Fisher’s tests were used to compare the differences between severe and critical clinical categories. Data are expressed as median (IQR - interquartile range) or number (% - percentages), and P < 0.05 was considered statistically significant.

| Confounding variables | Severe (Plasm) |  |  | Critical (Plasm) |  |  | Critical (BAL) |  |  |
| --- | --- | --- | --- | --- | --- | --- | --- | --- | --- |
| | non-GC | GC | P value (\$) | non-GC | GC | P value (\$) | non-GC | GC | P value (\$) |
| No | 6 | 10 |  | 8 | 8 |  | 3 | 17 |  |
| Age, median (IQR) | 75 (63-79) | 68 (55-76) | 0.415 | 65 (52-74) | 76 (60-79) | 0.316 | 74 (72-80) | 62 (53-76) | 0.243 |
| Gender, No (%) |  |  | 0.118 |  |  | 1.000 |  |  | 0.920 |
| Male | 1 (16.7) | 7 (70.0) |  | 6 (75.0) | 6 (75.0) |  | 2 (66.7) | 11 (64.7) |  |
| Female | 5 (83.3) | 3 (30.0) |  | 2 (25.0) | 2 (25.0) |  | 1 (33.3) | 6 (35.3) |  |
| Diabetes, No (%) | 4 (40.0) | 6 (60.0) | 1.000 | 1 (12.5) | 3 (37.5) | 0.569 | 1 (33.3) | 3 (17.6) | 0.450 |
| Hypertension, No (%) | 2 (20.0) | 8 (80.0) | 1.000 | 3 (37.5) | 5 (62.5) | 0.619 | 3 (100.0) | 6 (35.3) | 0.073 |
| Obesity, No (%) | 0 (0.0) | 1 (10.0) | 1.000 | 2 (25.0) | 2 (25.0) | 1.000 | 0 (0.0) | 3 (17.6) | 1.000 |

**Table S9.** Impact of Covid-19 confounding variables on the comparative analysis between high levels of lipid mediators in plasma and BAL samples from severe and critical patients. We used the same confounding variables associated with Covid-19 reported on Table S7. This analysis was based on plasma and BAL levels of lipid mediators (AA, 5-HETE, and 11-HETE) from severe/critical Covid-19 patients reported in Figure 4 (Panels: B, C, and D). None of the confounding variables tested had significant potential to interfere with the comparison between altered plasm and BAL levels of lipid mediators in severe/critical patients. §Mann-Whitney, chi-square, or Fisher’s tests were used to compare the differences between severe and critical patients. Data are expressed as median (IQR - interquartile range) or number (% - percentages), and P <0.05 was considered statistically significant.

| Confounding variables | Severe/Critical<br>(Plasm) | Severe/Critical<br>(BAL) | P value<br>(§) |
| --- | --- | --- | --- |
| No | 25 | 18 |  |
| Age, median (IQR) | 60 (53-77) | 64 (52-83) | 0.6395 |
| Gender, No. (%) Male |  |  | 0.1394 |
| Female | 14 (56.0) | 14 (77.8) |  |
| Diabetes, No. (%) | 11 (44.0) | 4 (22.2) |  |
| Hypertension, No. (%) | 7 (28.0) | 5 (27.8) | 0.9872 |
| Obesity, No. (%) | 16 (64.0) | 9 (50.0) | 0.3586 |
|  | 3 (12.0) | 4 (22.2) | 0.4274 |

## Supplementary Figures

**Figure S1.**
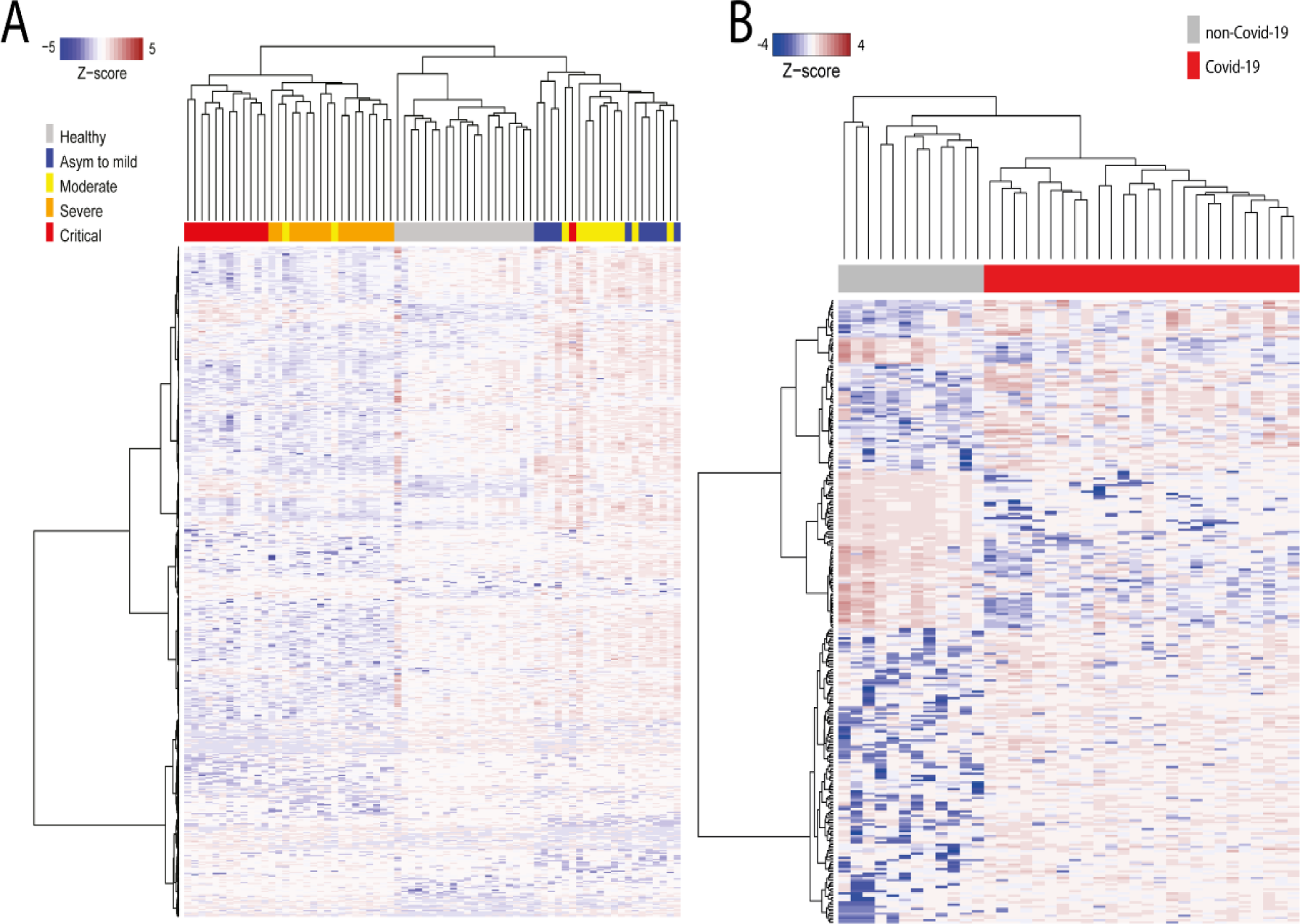
Metabolic signatures of plasma and BAL samples from patients infected or not with SARS-CoV-2. Hierarchical clustering based on different characteristics of abundant metabolites (ANOVA-FDR <0.1) between controls and patients positive for SARS-CoV-2 at different stages of the disease in plasma (A) and non-Covid-19 versus Covid-19 BAL patients (B). In (A), the clinical classification of Covid-19 was: healthy participants (n=20), asymptomatic-to-mild (n=10), moderate (n=12), severe (n=16), and critical (n=13) groups. In (B), the same analysis was performed with data from BAL samples of non-Covid-19 (n=12) and Covid-19 (n=26) patients. The detection levels of metabolites were defined using Z-score normalisation. The analysis of hierarchical clustering (A) shows highly different metabolomic profiles in the plasma for each group of individuals according to disease severity. BAL analysis (B) of samples from Covid-19 patients demonstrated an increase in the abundance of metabolites compared to non-Covid-19 individuals. The results indicated that SARS-CoV-2 infection induces changes in the metabolic profile of humans. Our data are in agreement with previous results showing virus-induced metabolic reprogramming in the host. The increased abundance of metabolites and their pathways, such as free fatty acids and amino acids, was correlated to the increase in viral proliferation, since these biomolecules can act as building blocks and fuel for this process ^141–143^.

**Figure S2.**
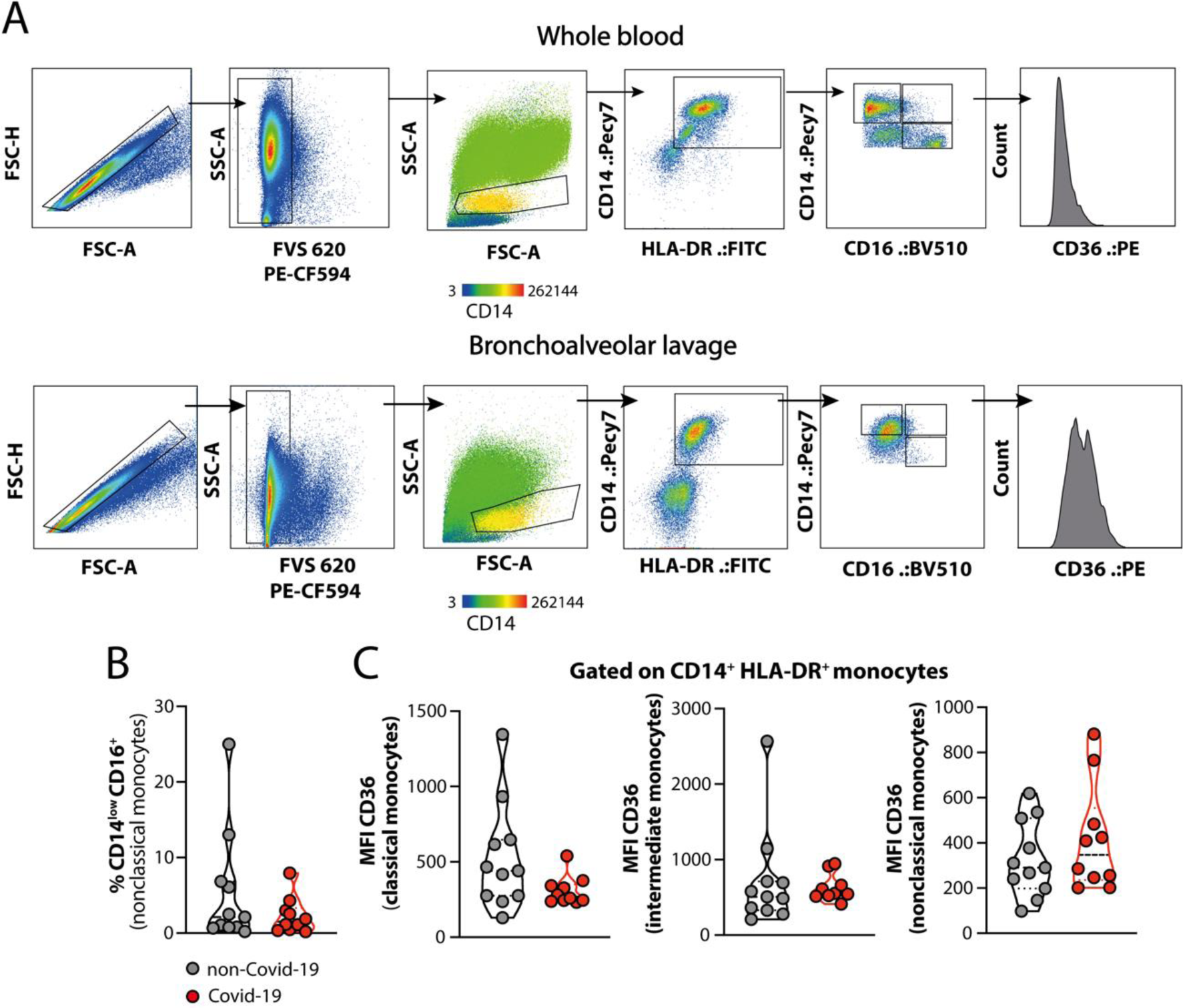
Gating strategy used for flow cytometry analysis of monocyte subsets and CD14/CD36 expression. In (A), dot plots show a representative gating strategy for the analysis of FSC-H/FSC-A, SSC-A/FVS-620 (viable cells), and SSC-A/FSC-A, followed by CD14^+^HLA-DR^+^ monocytes and the classical (CD14^high^CD16^-^), intermediate (CD14^+^CD16^+^) or non-classical CD14^low^CD16^+^ subsets, with subsequent CD36 mean fluorescence intensity (MFI) in whole blood (upper panel) and bronchoalveolar lavage fluid (BAL, bottom panel). In (B), a violin plot shows the frequency of non-classical monocytes from BAL of non-Covid-19 and Covid-19 patients. In (C), CD36 MFI in classical, intermediate, and non-classical monocytes from BAL of non-Covid-19 and Covid-19 samples. Differences between groups were calculated using the Mann-Whitney test. The membrane markers and gating strategies used to define the different monocyte subpopulations in the blood and BAL samples were adapted from a previous publication (ref). In BAL, there was a tendency to reduce non-classical monocytes in Covid-19 patients compared to non-Covid-19 patients (Figure 2SB), as well as a decrease in CD36 expression in classical and intermediate monocytes (Figure 2SC), although the difference was not statistically significant. Although not significantly, this reduction may have a biological importance. These data are similar to the profile observed in the blood monocytes of Covid-19 patients (Figure 3J).

**Figure S3.**
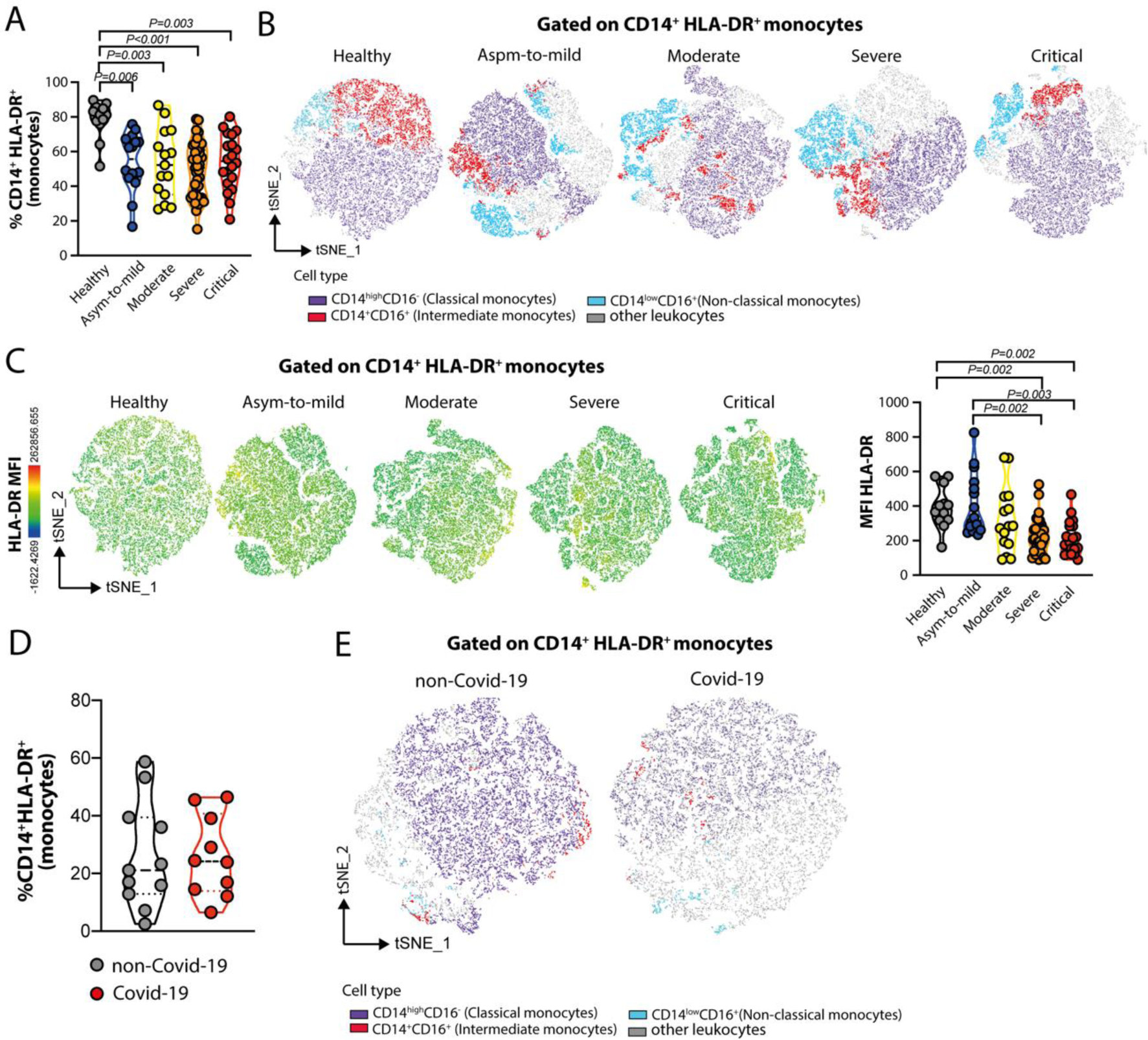
Determination of monocyte subsets in the blood and BAL samples from non-Covid-19 and Covid-19 patients. Peripheral blood from healthy participants (n=12), and asymptomatic-to-mild (n=15), moderate (n=15), severe (n=35), or critical (n=20) patients, and BAL from Covid-19 (n=10) and non-Covid-19 (n=11) participants were collected and used for the characterisation of monocyte subpopulations by flow cytometry and t-distributed stochastic neighbour embedding (t-SNE) analysis. (A) Frequency of CD14^+^HLA-DR^+^ cells. (B) t-SNE evaluation of classical (CD14^high^CD16^-^), intermediate (CD14^+^CD16^+^), and non-classical CD14^low^CD16^+^ blood monocytes in Covid-19 patients, categorised according to disease severity. (C) Colour mapping t-SNE and violin plot showing the mean fluorescence intensity (MFI) of HLA-DR expression in the circulating monocytes of Covid-19 subjects. In (D) and (E), samples of BAL from non-Covid-19 and Covid-19 patients were evaluated to determine the frequency of CD14^+^HLA-DR^+^ cells and classical, intermediate, or non-classical monocyte subsets, respectively. Differences among groups were calculated using the Kruskal-Wallis test with Dunn’s multiple comparison post-test, and the corresponding values are indicated in the figures. In comparison to healthy participants (Figure S3A, S3B, and 3SC), a significant reduction was observed in the intermediate monocyte population, as well as in the expression of HLA-DR molecules in the blood of Covid-19 patients, depending on disease severity. The diminishment of HLA-DR in monocytes correlated with TNF values obtained in the blood of patients positive for SARS-CoV-2 (Figure 2P; principal article), since monocytes with a pro-inflammatory profile are the main producers of this cytokine ^144^. Similarly, reduced intermediate monocytes were found in the BAL of patients positive for Covid-19 (Figure S3E). Interestingly, these patients also had a significant increase in the production of IL-8, IL-10, and IL-6 cytokines in the blood and BAL, as shown before (Figure 2M, 2N, and 2Q and Figure 4A; principal article). The high production of cytokines, especially IL-6, in patients with Covid-19, has been correlated with a decrease in the expression of HLA-DR in monocytes, while the therapeutic inhibition of IL-6 re-established the expression of this molecule in those individuals^145^

**Figure S4.**
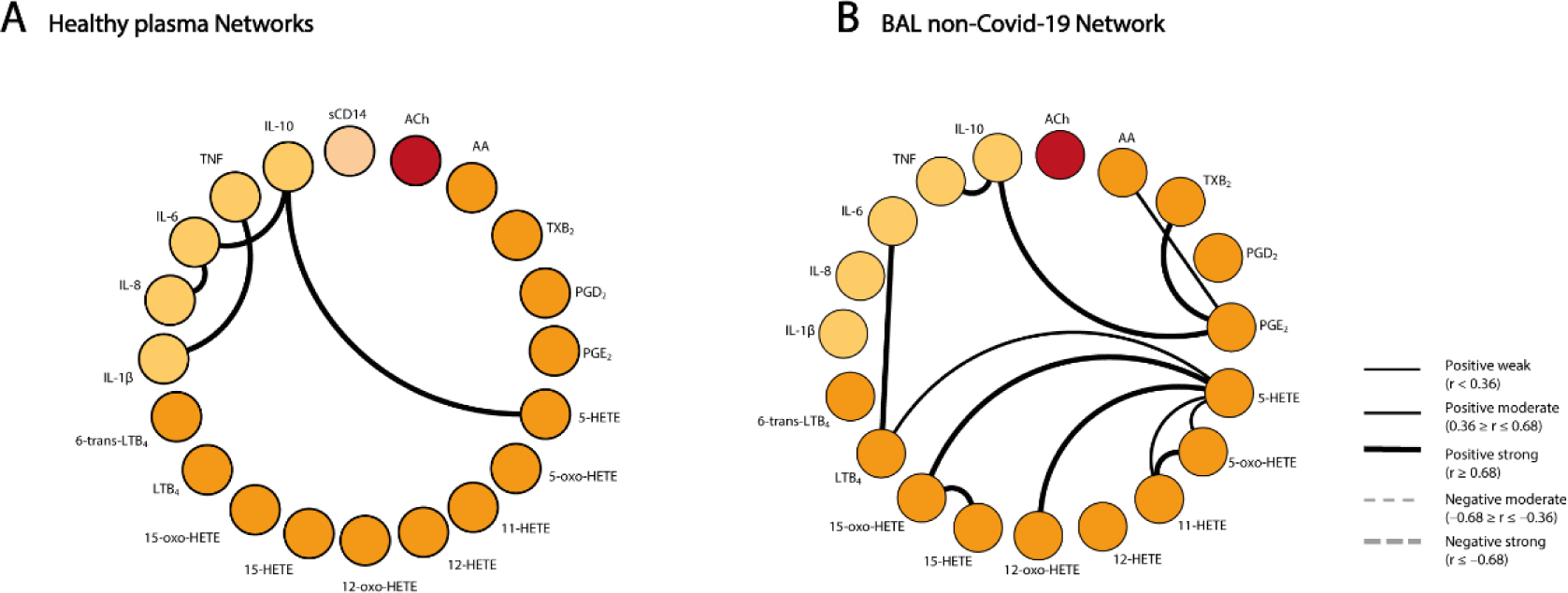
Biomarker networks in non-Covid-19 and healthy participants. A network of interactions between lipid mediators, ACh, sCD14, and cytokines in (A) healthy participants (n=39) and (B) BAL non-Covid-19 participants (n=13). Network layouts of personalised biomarkers were set up to identify the relevant association in healthy and non-Covid-19 participants. Each connecting line denotes a significant correlation between a pair of markers. Continuous lines represent positive correlations, while dashed lines represent negative correlations (*p*<0.05). The degree of significance is represented by the thickness of the line. Correlations were determined using Spearman’s test; the values of *r* and *p* were used to classify the connections as weak (*r* ≤ 0.35, *p*<0.05), moderate (*r* = 0.36–0.67, *p*<0.01), or strong (*r* ≥ 0.68, *p*< 0.001). The absence of a line indicates the non-existence of the relationship. To evaluate the relationship between levels of lipid mediators, ACh, sCD14, and cytokines in non-Covid-19, a series of correlation analyses were performed. When comparing the interactions analysed between the molecules of the healthy and non-Covid-19 groups, we observed that (A) In the blood of healthy individuals, the interactions were found to occur mainly between cytokines (IL-1β, TNF, IL-8, IL-6, and IL-10) and the lipid mediator 5-HETE. (B) In thee BAL samples from hospitalised non-Covid-19 patients, lipid mediators were found to mediate these interactions and influence the clinical outcomes of this group of patients.

**Figure S5.**
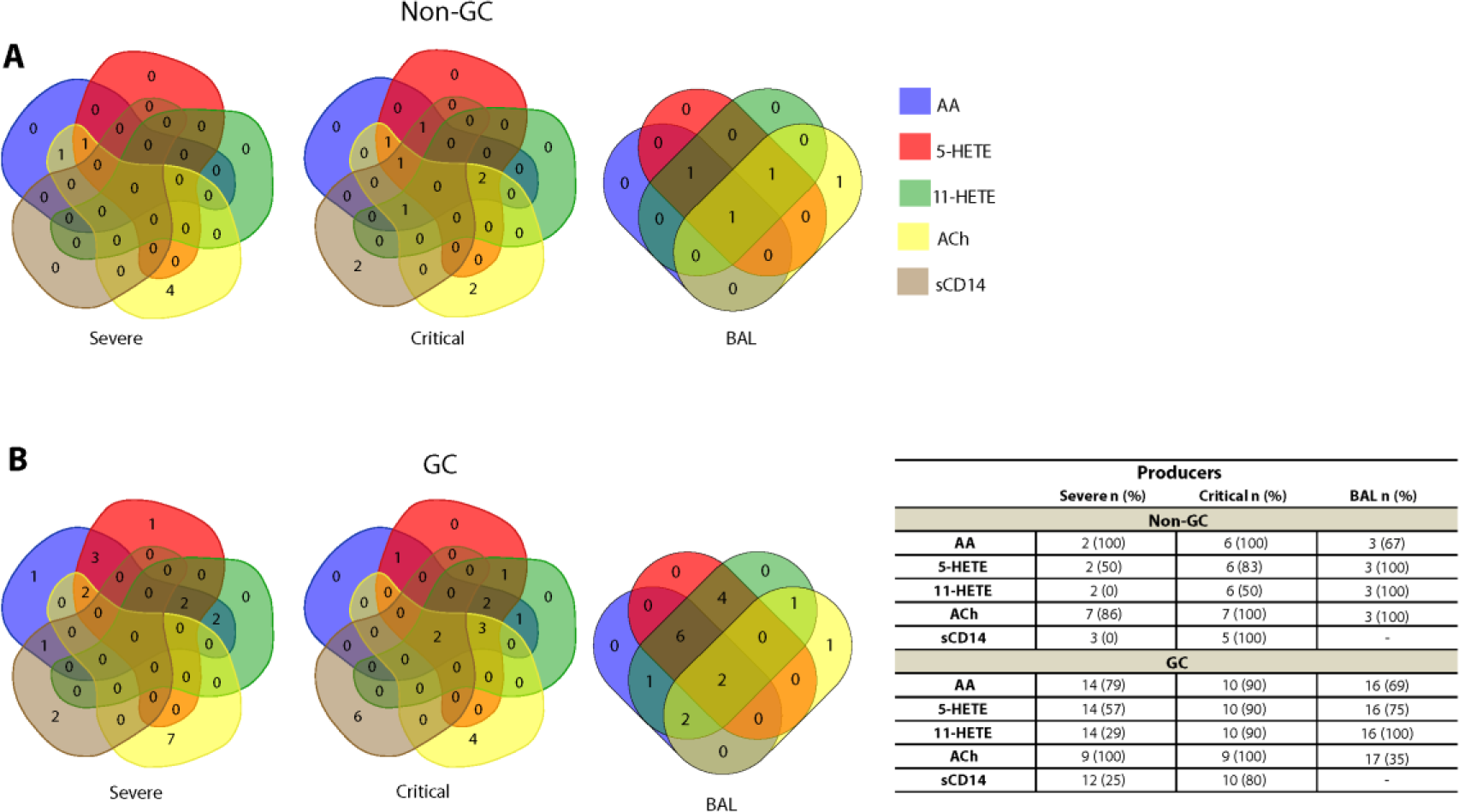
Treatment of severe and critical SARS-CoV-2-infected patients with glucocorticoids inhibits ACh release. The production of lipid mediators and the shedding of sCD14 in patients with Covid-19 at the severe and critical stages was not modified by treatment with glucocorticoids (GC). However, a marked reduction was observed in ACh release. Intersection analysis depicted in Venn diagrams revealed the number of patients producing AA, 5-HETE, 11-HETE, ACh, or sCD14, above the control (healthy participants) values, in the absence (A) or presence (B) of GC therapy. The table shows the number (*n*) of plasma or BAL samples from Covid-19 patients at the severe and critical stages of disease, for each mediator. The percentages shown in parentheses represent the frequency of patients producing the respective mediator for the corresponding sample number. The Venn diagrams show the intersections of AA, 5-HETE, 11-HETE, ACh, and sCD14s in Covid-19 patients at the severe and critical stages of the disease, in both blood and BAL. No significant changes were observed in the quantification of lipid mediators or sCD14, comparing patients treated with or without glucocorticoids (Figure S5A and FigureS5B). Interestingly, critically ill patients treated with glucocorticoids showed a 20% and 65% reduction in the ACh concentration in both blood and BAL samples, respectively, compared to patients who were not administered drugs (Figure S5B). While a decrease in free-AA and its metabolites (5-HETE and 11-HETE) has not been reported in the literature, some studies have demonstrated the beneficial effects of the use of glucocorticoids in other respiratory syndromes, having been found to reduce the production of AA-derived mediators ^72, 146^. In agreement with our results, there is no evidence to support corticoid treatment for Covid-19 ^147^. Data related to the effect of glucocorticoids on ACh release in viral infections are scarce. However, according to our previous findings on scorpion envenomation^3^, glucocorticoid treatment should be initiated early after infection to block the release of lipid mediators, which could lead to the inhibition of premature ACh release. On the contrary, the late administration of GC could place patients at a point of no return, with early ACh release impacting vital organs, damaged by the negative impacts of this neurotransmitter before GC administration. Moreover, it should be considered that, in Covid-19, other unknown mechanisms may control ACh release, in addition to the COX-2 dependent metabolites.

**Figure S6.**
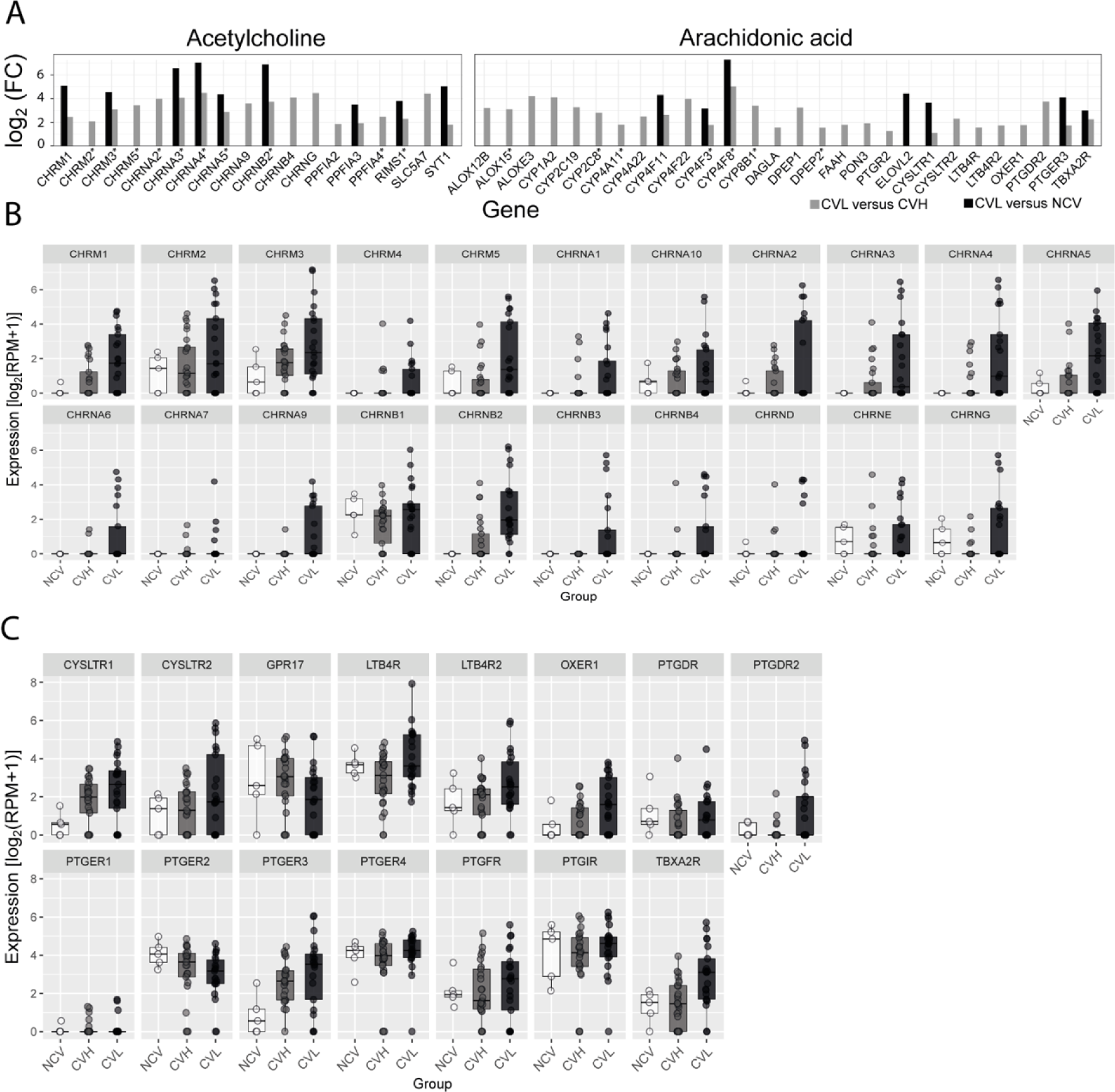
Differential expression and profile of genes involved in AA and ACh pathways from SARS-CoV-2 deceased patients. Abbreviations: CVL, Covid-19 low viral load; CVH, Covid-19 high viral load; NCV, non-Covid-19 viral load ^9, 50, 110, 113^. As shown in (A), DEGs were upregulated in CVL versus CVH patients and CVL versus NCV patients. The transcript expression of cholinergic receptors (muscarinic and nicotinic) (B) and eicosanoid receptors (C) was normalised in the NCV, CVH, and CVL groups. CVL patients displayed increased pulmonary levels of expression of genes associated with the ACh and AA pathways, encoding for cholinergic receptors, the ACh release cycle, AA metabolism, and eicosanoid receptors, compared to CVH or NCV patients. The expression profile of some genes may favour inflammatory events, such as upregulated cholinergic and eicosanoid receptors (CHRM3, CHRNA3, CHRNA5, and OXER1) and low levels of the anti-inflammatory cholinergic receptor (CHRNA7)^9, 50, 110, 113^

## Notes

### Competing Interest Statement

The authors have declared no competing interest.

### Clinical Trial

This is an observational study.

### Author Declarations

The entire study was approved by two Brazilian ethics oversight body: i) a local Committee, Comitê de ética em Pesquisa from Faculdade de Ciências Farmacêuticas de Ribeiréo Preto of the Universidade de Séo Paulo (CEP-FCFRP-USP - https://fcfrp.usp.br/en/institucional/comissoes/comite-etica-pesquisa/ -), coordinated by Prof. Dr. Cleni Mara Marzocchi Machado, Ph.D.; ii) a national Committee, Conselho Nacional de Pesquisa em Humanos (CONEP - https://plataformabrasil.saude.gov.br/login.jsf -), coordinated by Jorge Alves de Almeida Venâncio, M.D., Ph.D. The research protocol (first approval document) and four amendments were approved and received the certificate of Presentation and Ethical Appreciation (CAAE: 30525920.7.0000.5403). We are sending an Ethical Approval Statement as a supplemental material, please see enclosed.

