## Supplementary material for "Cholinergic and lipid mediators crosstalk in Covid-19 and the impact of glucocorticoid therapy": ETHICAL APPROVAL STATEMENT

UNIVERSIDADE DE SÃO PAULO
Faculdade de Ciências Farmacêuticas de Ribeirão Preto
Comitê de Ética em Pesquisa / Research Ethics Committee

Ribeirão Preto, January 3, 2021.

**Subject: Approval by the Research Ethics Committee**

To
Prof. Dr. Cristina Cardoso Ribeiro de Barros
School of Pharmaceutical Sciences of Ribeirão Preto, University of São Paulo (FCFRP-USP)
Department of Clinical Analyses, Toxicology and Food Sciences

Dear Professor,

We inform that the research project entitled **"Prospective evaluation of gene expression and humoral response in severe COVID-19: search for potential biomarkers for disease evolution in patients infected with SARS-COV2"**, was approved by the Research Ethics Committee (CEP) of the FCFRP-USP and the National Research Ethics Commission (CONEP), which constitute the Brazilian body for ethical supervision in research involving human beings (CEP / CONEP system), according to Resolution No. 466/2012 of the National Health Council, Ministry of Health, Brazil. Resolution 466/2012 provides the guidelines and regulatory standards for research involving human beings in Brazil, which is based on the main documents that constitute the pillars of recognition and affirmation of the dignity, freedom and autonomy of human beings, such as Helsinki Declaration of 1964 and its updates; the 2004 Universal Declaration on Bioethics and Human Rights; and the Federal Constitution of the Federative Republic of Brazil, whose objectives are consistent with international documents on ethics, human rights and development.

The present research protocol is registered in the CEP / CONEP system under the number of Certificate of Presentation of Ethical Appreciation (CAAE) 30525920.7.0000.5403. The experimental design / current methods of this research protocol approved by the CEP / CONEP system are described below. Then, the history of the ethical analysis since the first submission of the research protocol and its respective amendments is presented.

*"Design/Methods: This study aims to investigate factors of susceptibility and resistance, as well as biomarkers of the disease caused by the SARS-COV2 virus, called COVID-19. For that, healthy participants from the USP community (Campus Ribeirão Preto), individuals with a positive molecular test for SARS-COV2 performed via Basic Health Units in the city of Ribeirão Preto and who are asymptomatic or have mild symptoms, as well as convalescents and patients will be selected from private hospitals (Hospital São Paulo and Hospital Unimed Ribeirão Preto) or another philanthropic hospital (Hospital Santa Casa de Misericórdia de Ribeirão Preto), in the same city, which will be evaluated clinically and laboratory, for the diagnosis of the disease, by the doctors responsible for the care. Men and women will be included, classified into control and sick groups, according to laboratory diagnosis and severity of COVID-19. After this categorization, the volunteers (100 per group) will be divided into healthy individuals with negative molecular*

*diagnosis for SARS-COV2 (group 1 - G1), asymptomatic or oligosymptomatic with positive test for the virus (group 2 - G2), hospitalized with moderate symptoms. positive diagnosis (group 3 - G3) and severe hospitalized symptoms, also with positive molecular test for SARS-COV2 (group 4 - G4), severe hospitalized symptoms, with negative molecular test for SARS-COV2 (group 6 - G6). In addition to convalescents, previously tested and confirmed positive for the virus, however in the current absence of signs or symptoms of COVID-19 (group 5 - G5). After the consent and signature of the Informed Consent Form (ICF) by the patients or guardians themselves, samples of venous blood, saliva or pulmonary fluid aspirate will be collected for laboratory processing related to this research project (some control participants will also have collection nasopharyngeal swap for molecular diagnosis of virus infection). Samples from participants in groups G1, G2 and G5 will be collected only once, for research. Patients in groups G3 and G4, will have blood, saliva (only G3) and / or pulmonary fluid aspirate (G4 and G6) collected at the time of admission to the hospital and after 48 hours, repeating the collection once every 7 days, until the end of the hospitalization period. The blood samples will be centrifuged to obtain plasma and leukocytes (buffy coat). The obtained cells will be immediately frozen in Trizol, for subsequent RNA extraction and transcriptomic analysis of gene expression of receptors, transcription factors, steroidogenic enzymes, cytokines, and other mediators related to the immune response. In plasma, cytokines will be quantified by multiplex assay, in addition to eicosanoids, steroid hormones, sphingolipids and ceramides, by mass spectrometry. In addition, the profiles of N-glycans in the Fcs of IgGs and anti-carbohydrate antibodies will be evaluated, to ascertain their correlation with the clinical outcomes of SARS-COV2 infections. New biotechnological tools that allow the study of the relationships between the virus, its cellular targets and antibodies will be produced. With these results together, it is expected to understand more deeply the pathophysiology of COVID-19, as well as to establish markers of disease evolution and morbidity".*

History of the approval ethical by CEP / CONEP system:

First approval: - CEP, April 6, 2020 - consubstanced opinion number 3,956,413; and CONEP, April 30, 2020 - consubstanced opinion number. 3,999,280;

Amendment 1 - approval: CEP, June, 8, 2020 - consubstanced opinion number 4,076,158; and CONEP, June 23,2020 - consubstanced opinion number 4,104,192;

Amendment 2 - approval: CEP, July 7, 2020 - consubstanced opinion number 4,134,068; and CONEP, July 28, 2020 – 4,176,358;

Amendment 3 - approval: CEP, November 19, 2020 - consubstanced opinion number 4,409,566; and CONEP, December 02, 2020 – 4,432,873;

Amendment 4 - approval: CEP, December 10, 2020 - consubstanced opinion number 4,454,975; and CONEP, December 15, 2020 - consubstanced opinion number 4,465,002.

The requirements for the ethical approval of this research protocol also included declarations of agreement from all health institutions (hospitals and basic health units), where the research participants will be recruited. The consultation of the patient's medical record is explained in the Informed Consent Form.

It is the responsibility of the researchers, according to Resolution 466/2012, item IV.5, letter d, to consider that the Informed Consent Form must be presented in two original documents, initialed in all its pages, and signed, at the end, by the research participant, or by his / her legal representative, as well as by the responsible researcher, or by the person (s) delegated by him / her, and the signature pages must be on the same sheet. The addresses and telephone or other contact, of those responsible for the research and of this Ethics Committee must be included.

The final report of the research must be sent to the CEP of the FCFRP-USP in its own form, as well as any changes, complications or interruptions, such as adverse events and eventual modifications to the protocol or team members, through the interposition of amendments on the Brazil Platform, which is the national and unified base of research records involving human beings for the entire CEP / CONEP system.

Prof. Dr. Cleni Mara Marzocchi Machado
Coordinator of the Research Ethics Committee of FCFRP-USP
